# Generalizable Long COVID Subtypes: Findings from the NIH N3C and RECOVER Programs

**DOI:** 10.1101/2022.05.24.22275398

**Authors:** Justin T. Reese, Hannah Blau, Timothy Bergquist, Johanna J. Loomba, Tiffany Callahan, Bryan Laraway, Corneliu Antonescu, Elena Casiraghi, Ben Coleman, Michael Gargano, Kenneth J. Wilkins, Luca Cappelletti, Tommaso Fontana, Nariman Ammar, Blessy Antony, T. M. Murali, Guy Karlebach, Julie A McMurry, Andrew Williams, Richard Moffitt, Jineta Banerjee, Anthony E. Solomonides, Hannah Davis, Kristin Kostka, Giorgio Valentini, David Sahner, Christopher G. Chute, Charisse Madlock-Brown, Melissa A Haendel, Peter N. Robinson, the N3C consortium, the RECOVER Consortium

## Abstract

Accurate stratification of patients with post-acute sequelae of SARS-CoV-2 infection (PASC, or long COVID) would allow precision clinical management strategies. However, the natural history of long COVID is incompletely understood and characterized by an extremely wide range of manifestations that are difficult to analyze computationally. In addition, the generalizability of machine learning classification of COVID-19 clinical outcomes has rarely been tested. We present a method for computationally modeling PASC phenotype data based on electronic healthcare records (EHRs) and for assessing pairwise phenotypic similarity between patients using semantic similarity. Our approach defines a nonlinear similarity function that maps from a feature space of phenotypic abnormalities to a matrix of pairwise patient similarity that can be clustered using unsupervised machine learning procedures. Using k-means clustering of this similarity matrix, we found six distinct clusters of PASC patients, each with distinct profiles of phenotypic abnormalities. There was a significant association of cluster membership with a range of pre-existing conditions and with measures of severity during acute COVID-19. Two of the clusters were associated with severe manifestations and displayed increased mortality. We assigned new patients from other healthcare centers to one of the six clusters on the basis of maximum semantic similarity to the original patients. We show that the identified clusters were generalizable across different hospital systems and that the increased mortality rate was consistently observed in two of the clusters. Semantic phenotypic clustering can provide a foundation for assigning patients to stratified subgroups for natural history or therapy studies on PASC.

## INTRODUCTION

Hundreds of millions of cases of acute Coronavirus disease 2019 (COVID-19) have been recorded since the beginning of the pandemic, and more than six million deaths had been reported by the World Health Organization by the end of March, 2022 *(1)*. The clinical presentation of COVID-19 ranges from asymptomatic infection to fatal disease, with many patients continuing to have heterogeneous, long-term, multi-system symptoms including fatigue, post-exertional malaise, dyspnea, cough, chest pain, palpitations, headache, arthralgia, weakness (asthenia), paresthesias, diarrhea, alopecia, rash, impaired balance, and memory or cognitive dysfunction *(2, 3)*. Although there is still no detailed and widely accepted case definition, post-acute sequelae of SARS-CoV-2 infection (PASC, long-haul COVID or long COVID) generally refers to a range of persistent or new symptoms beyond three or four weeks of the initial infection *(4–6)*. The NIH REsearching COVID to Enhance Recovery (RECOVER) Initiative program defines PASC as ongoing, relapsing, or new symptoms, or other health effects occurring after the acute phase of SARS-CoV-2 infection (i.e., present four or more weeks after the acute infection). The World Health Organization (WHO) has developed a case definition of “post COVID-19 condition” suggesting that the syndrome is usually diagnosed several months after the onset of acute symptoms of COVID-19 based on new-onset or lingering symptoms (e.g., fatigue, dyspnea, cognitive dysfunction) which cannot be explained by an alternative etiology and which continue for at least two months *(7)*. In this work, we will use the term long COVID to refer to patients given a diagnosis using the newly introduced ICD-10 U09.9 code (“Post COVID-19 condition”), acknowledging that the code may be applied inconsistently given the lack of a gold standard definition..

The pathogenesis of long COVID is incompletely understood, but it appears likely that different pathogenetic mechanisms or combinations thereof may drive disease in individual patients. Potential factors that may contribute to the development of long COVID include aberrant immune responses, persistent viral replication, redox imbalance, formation of fibrinolysis-resistant amyloid fibrin microclots, and consequences from acute SARS-CoV-2 injury to one or multiple organs *(8–17)*. At present, there is no specific treatment for long COVID and it is imperative to achieve a better understanding of long COVID subtypes.

Our understanding of the natural history of long COVID is still incomplete. Limited emerging evidence suggests the existence of clinical subtypes or clusters characterized by the predominance of symptoms such as pain or cardiovascular manifestations, or by a paucity of symptoms *(18)*. However, computational methods to characterize long COVID subtypes based on comprehensive phenotypic analysis are lacking, as are approaches to assess the generalizability of the resulting clusters across different patient cohorts. In this study, we constructed a cohort of 2464 patients diagnosed with long COVID using the newly introduced ICD-10 U09.9 code (“Post COVID-19 condition”) from multicenter electronic health record (EHR) data available through the National COVID Cohort Collaborative (N3C), a harmonized EHR repository with 2,909,292 COVID-19 positive patients as of March 16, 2022. Previous work mapped 287 unique clinical findings previously reported in studies of long COVID *(19)* to the Human Phenotype Ontology (HPO), which is widely used to support differential diagnosis and translational research in human genetics *(20, 21)*. Here, we introduce an approach that calculates the semantic similarity between patients by transforming EHR data to phenotypic profiles using the HPO. The method identifies distinct clusters of long COVID patients that show highly significant correlations with pre-existing conditions and generalize across different hospital systems.

## RESULTS

### A cohort of patients diagnosed with PASC

As of March 16, 2022, the N3C platform (“Enclave”) contained data for 2,909,292 patients diagnosed with acute COVID-19, and 21 data partners had begun to use the newly introduced ICD-10 diagnosis code U09.9 for Post COVID-19 condition, providing data for 5,645 patients with this diagnosis (Figure 1). Phenotypic features observed in the post-acute COVID-19 period were mapped from OMOP codes to HPO terms. The post-acute COVID-19 period was defined as starting 21 days after the earliest COVID-19 index date for outpatients, and 21 days after the end of hospitalization for inpatients. The COVID-19 index date for each patient was defined as the earliest date of any positive PCR or antigen SARS-CoV-2 test or COVID-19 U07.1 diagnosis.

**Figure 1.**
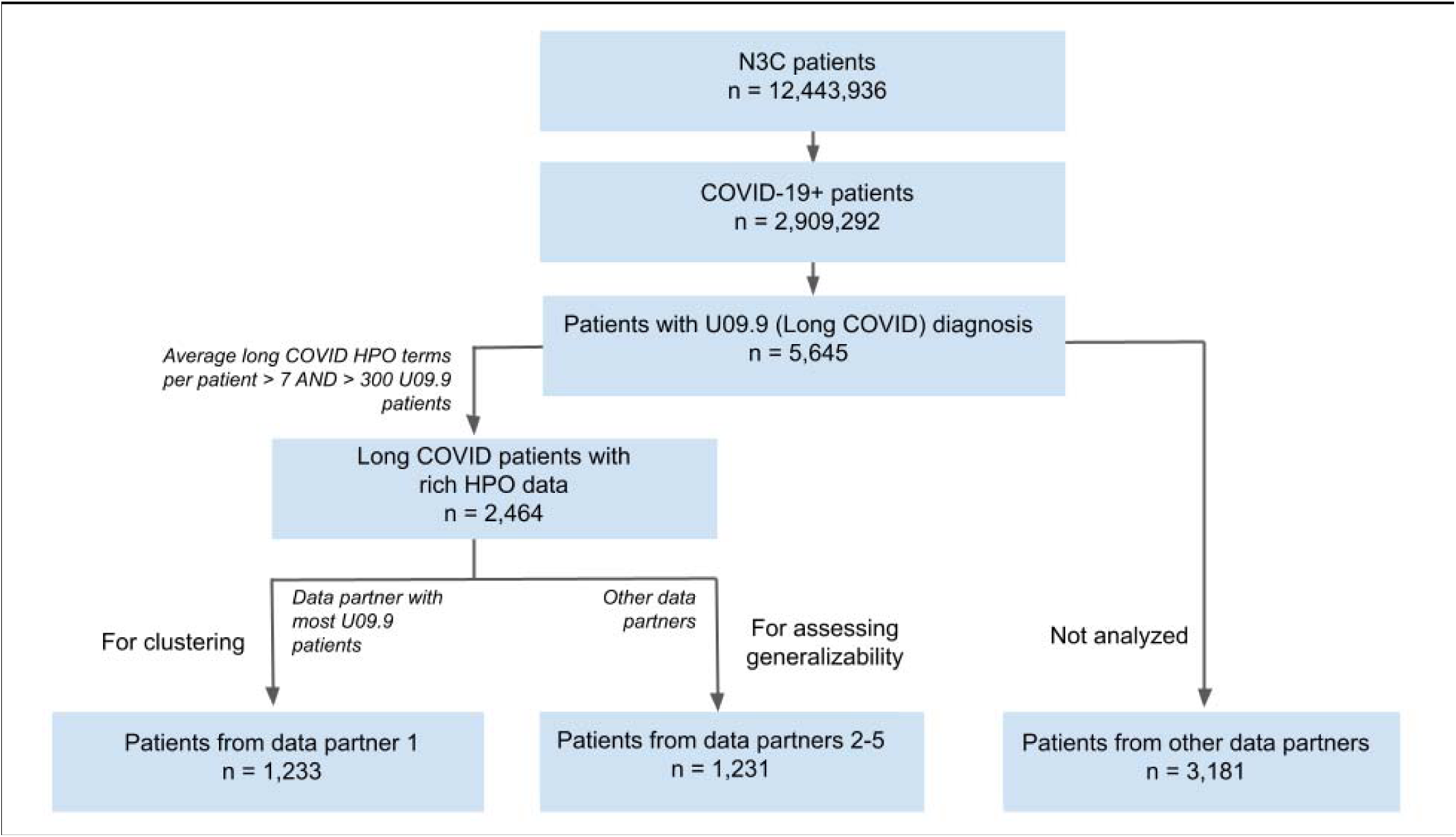
Cohort construction. Patients with long COVID (U09.9 diagnosis) were extracted from the much larger dataset of the N3C. Long COVID patients were selected from the five data partners that provided data for at least 300 U09.9 patients and had an average of at least 7 long COVID HPO terms per patient. The data partner with the most U09.9 patients (data partner 1) was chosen for clustering, and additional U09.9 patients from four other data partners (data partners 2-5) were chosen to assess generalizability.

### Phenotypic Clustering of Patients with long COVID

We hypothesized that consistent subgroups of patients with long COVID can be defined based on the spectrum of phenotypic features in the patients’ electronic health records (EHR). Our previous analysis identified 287 clinical findings previously reported in studies on long COVID and coded these findings using terms of the Human Phenotype Ontology (HPO) *(19, 21)*. Numerous algorithms have been developed that define a fuzzy, specificity-weighted similarity metric between a patient and a computational disease model or between pairs of patients *(22–25)*. Here, we adapted an algorithm called Phenomizer that calculates semantic similarity between a pair of patients based on phenotypes (Methods) *(26)*. Common clustering methods define feature vectors with one field for each measured quantity. In principle, one could define a feature vector with 287 dimensions, one for each of the clinical findings related to long COVID, and for each clinical finding identified in a patient, a “1” would be placed in the corresponding field of the vector, otherwise a “0”. Patient similarity could then be measured by calculating the cosine between any two such vectors, which essentially counts the number of exact matches normalized by the total number of features in each vector. This procedure would not capture the fact that some features are similar. For instance, although dyspnea and hypoxemia are both abnormalities of respiratory physiology, they are represented by different fields in the feature vector and thus if one patient was recorded to have dyspnea and another hypoxemia, this would not contribute to the similarity score. Another drawback to a simple 0/1 feature vector for the 287 clinical findings would be that matches between more or less specific findings would be weighted equally. The Phenomizer algorithm uses the structure of the ontological hierarchy to identify partial matches between related clinical findings, and it leverages the information content of each term, which is a measure of specificity, to weight the matches. The Phenomizer is thus a nonlinear mapping from the original feature space of clinical findings to a pairwise similarity matrix that implements a fuzzy, specificity-weighted matching strategy. The resulting similarity matrix can be used as input to a number of clustering algorithms (Figure 2).

**Figure 2.**
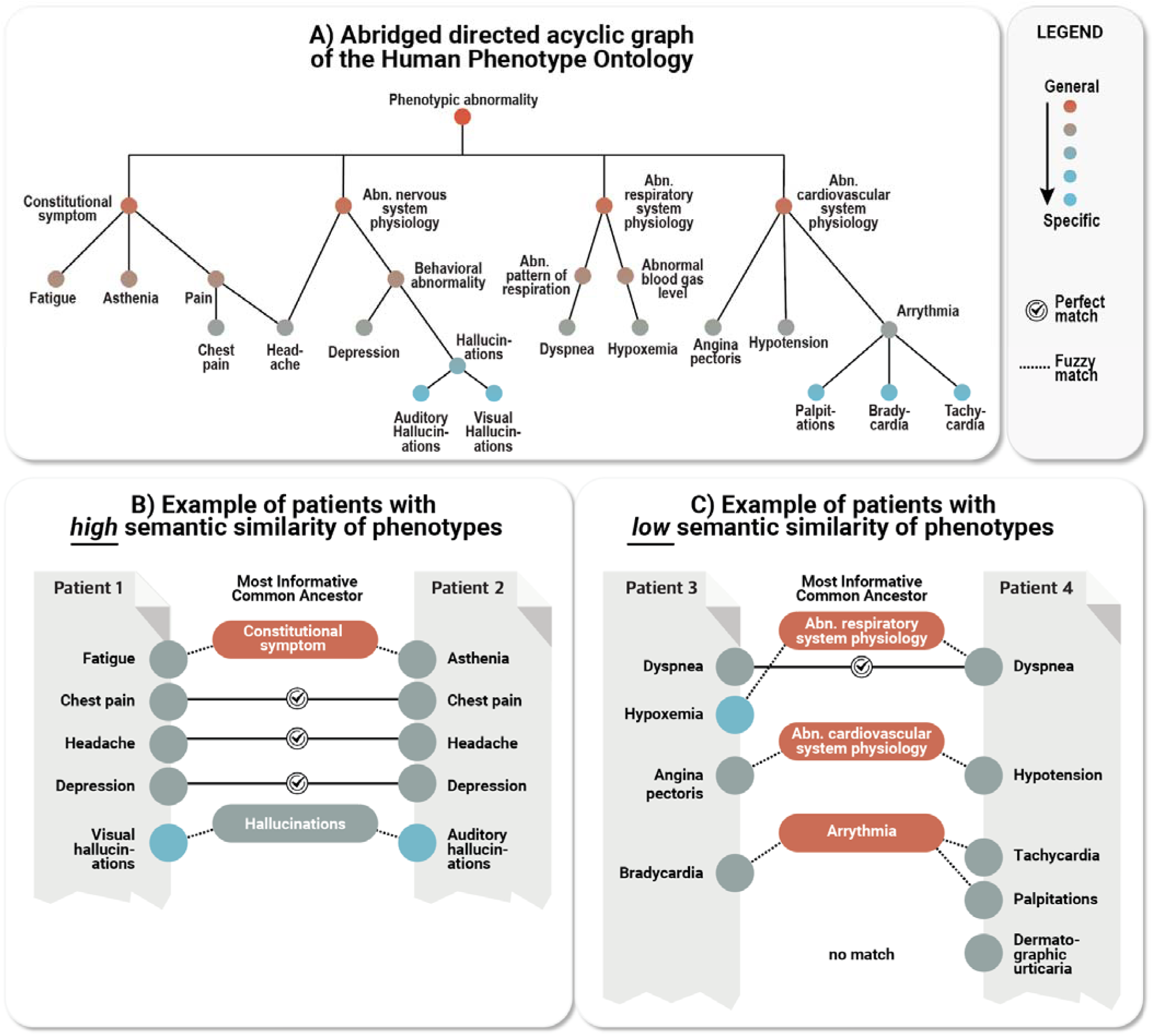
Calculating patient semantic similarity based on HPO phenotypes. **A)** HPO terms are arranged in a directed acyclic graph with specific terms such as *Bradycardia* (HP:0001662) being related to more general terms (here: *Arrhythmia*; HP:0011675) by subtype relations. An excerpt of the entire ontology (15,247 terms) is shown. **B)** Example showing a pair of patients with relatively high phenotypic similarity; for each of the HPO terms in patient 1, the best match is sought in patient 2. If an exact match is not found, the algorithm searches for the most informative common ancestor (MICA) in the ontology; the information content (a measure of specificity) of the exact matching term or most specific ancestor term is calculated to determine the specificity. For instance, *Visual hallucinations* (HP:0002367) and *Auditory hallucinations* (HP:0008765) are not an exact match, so the information content of their MICA *Hallucinations* (HP:0000738) is chosen. *Hallucinations* (HP:0002367) is still relatively specific (and shown in gray), while the MICA of *Angina pectoris* (HP:0001681) and *Hypotension* (HP:0002615) is more general (shown in red) and contributes less to the matching score. **C)** Example of a pair of patients with a relatively lower similarity due to (specific) fewer exact matches and one unmatched term. The pairwise similarity is calculated in this way for all pairs of patients to construct the similarity matrix that is used for clustering (Figure 3).

To leverage this procedure for analysis of N3C data, we mapped the 287 long COVID-associated HPO terms*(19)* to corresponding Observational Medical Outcomes Partnership (OMOP) codes *(27)* (see Methods). Of these, 116 terms were identified in the data (Supplemental Tables S1-S11). The terms not found in the data largely were clinical or patient-reported features that are not commonly represented in EHR data, such as *Centrilobular ground-glass opacification on pulmonary HRCT* (HP:0025180) or *Ocular pruritus* (HP:0033841), and were not included in further analyses.

We selected data partners that provided at least 300 U09.9 patients and an average of at least seven HPO terms per patient (Figure 1). This threshold was chosen to include data partners with a sufficient number of patients with a sufficient depth of phenotypic information available in EHR data to assess patient similarity. For clustering, we selected U09.9 patients from the data partner (referred to here as data partner 1, as data regulations disallow use of real data partner names or IDs) that supplied data for the greatest number of U09.9 patients (1233 patients). For assessment of the generalizability of the clusters to other data partners, we selected the remaining U09.9 patients from the remaining data partners (referred to here as data partners 2-5, again due to data regulations) (1,231 patients). We calculated the frequency with which each term was used in the total group of 1233 patients from data partner 1 and used this value to determine the information content (a measure of specificity; see Methods) for each term.

In order to calculate pairwise phenotypic similarity of patients at data partner 1 for clustering, we leveraged the Phenomizer algorithm to calculate a 1233 × 1233 similarity matrix for the 1233 patients at data partner 1. K-means clustering was applied to the data and the number of clusters was determined to be 6 based on visual inspection of the ‘elbow’ curve (Figure 3; Supplemental Figure 1). We note that although the determination of cluster number by this method is subjective, the major findings were similar with 4 or 5 clusters (Supplemental Figure S2-S3).

**Figure 3.**
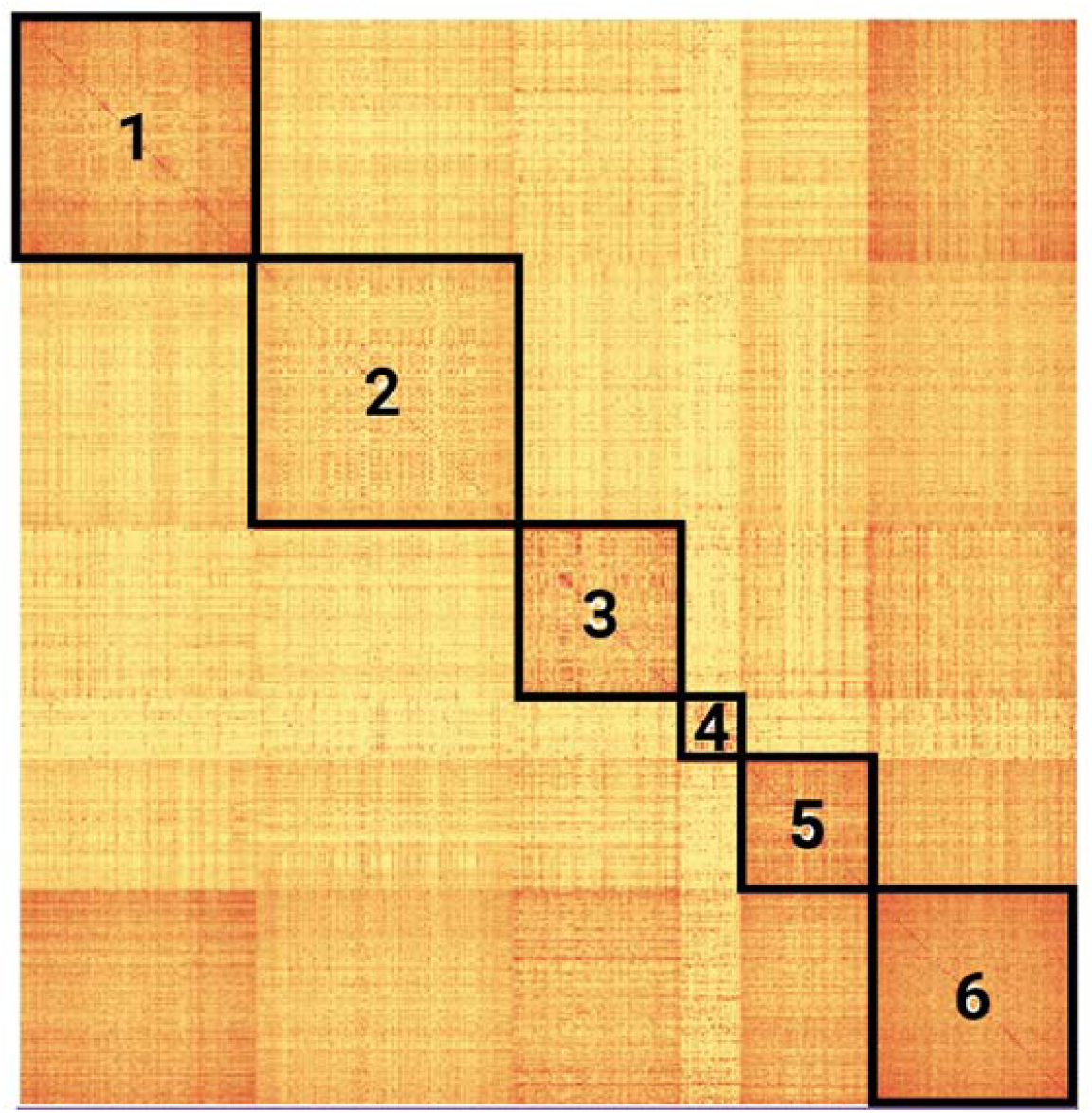
Patient similarity matrix illustrating long COVID subtypes in data partner 1. A heatmap representing the 6 clusters created by k-means clustering is shown. Cluster hierarchy was calculated using the nearest point algorithm and Euclidean distance.

### Characterization of PASC Clusters

We characterized the features of each of the six clusters with respect to age, gender, and race/ethnicity (Table 1). The six clusters contained between 70 and 301 patients, and differed significantly with respect to rate of hospitalization, age, gender, and ethnicity. Patients in clusters 1 and 6 were overall older, more likely to have been hospitalized during their acute COVID-19 infection, more likely to be male, and were less likely to be of White non-Hispanic race/ethnicity. Patients in clusters 3, 4, and 5 were almost entirely non-hospitalized, younger, and more likely to be female.

**Table 1.** Characteristics of the study population in data partner 1. For the overall study population and for each cluster, age, gender, and race/ethnicity are shown. Data for characteristics for which there were fewer than 20 patients, and data about race/ethnicities for which there were fewer than 20 patients overall (Other Non-Hispanic, Native Hawaiian or Other Pacific Islander Non-Hispanic, Asian Non-Hispanic) are not shown to reduce the risk of patient re-identification. **p < 0.001, *p < 0.05 by one-way ANOVA (age) or chi squared test (all others).

|  | Overall | Cluster 1 | Cluster 2 | Cluster 3 | Cluster 4 | Cluster 5 | Cluster 6 |
| --- | --- | --- | --- | --- | --- | --- | --- |
| n | 1233 | 276 | 301 | 195 | 70 | 148 | 243 |
| Acute COVID-19 Inpatient** | 424 (34.6%) | 203 (74.1%) | 21 (7.0%) | <20 | 0 | <20 | 170 (70.0%) |
| age - mean (SD)** | 51.9 (16.5) | 58.7 (17.6) | 50.0 (15.3) | 48.5 (15.2) | 47.0 (16.4) | 44.6 (13.4) | 55.0 (16.3) |
| Female** | 714 (58.2%) | 112 (40.9%) | 182 (60.7%) | 127 (65.5%) | 48 (69.6%) | 104 (70.7%) | 141 (58.0%) |
| Black or African American Non-Hispanic | 60 (4.9%) | <20 | <20 | <20 | <20 | <20 | <20 |
| White Non-Hispanic* | 882 (71.9%) | 186 (67.9%) | 228 (76.0%) | 153 (78.9%) | 54 (78.3%) | 107 (72.8%) | 154 (63.4%) |
| Hispanic or Latino Any Race | 202 (16.5%) | 52 (19.0%) | 42 (14.0%) | <20 | <20 | 26 (17.7%) | 53 (21.8%) |
| Unknown race/ethnicity | 58 (4.7%) | <20 | <20 | <20 | <20 | <20 | <20 |

To further characterize each of the six clusters, we identified HPO terms that tended to occur among patients in certain clusters (Figure 4). Of the 287 HPO terms we identified as being used in published cohort studies on long COVID *(19)*, only 116 were identified in our data. The presence or absence of each of the 116 HPO terms used for clustering was treated as a categorical variable whose distribution among the six clusters was assessed using a chi-squared test. Of the 116 HPO terms that were tested, 63 were significantly correlated with cluster membership following Bonferroni correction. Of these, 26 terms had a corrected p-value of less than 10^5^ and were present in at least 20% of patients in one or more clusters and were therefore considered to be the characteristic features that best defined the clustering.

**Figure 4.**
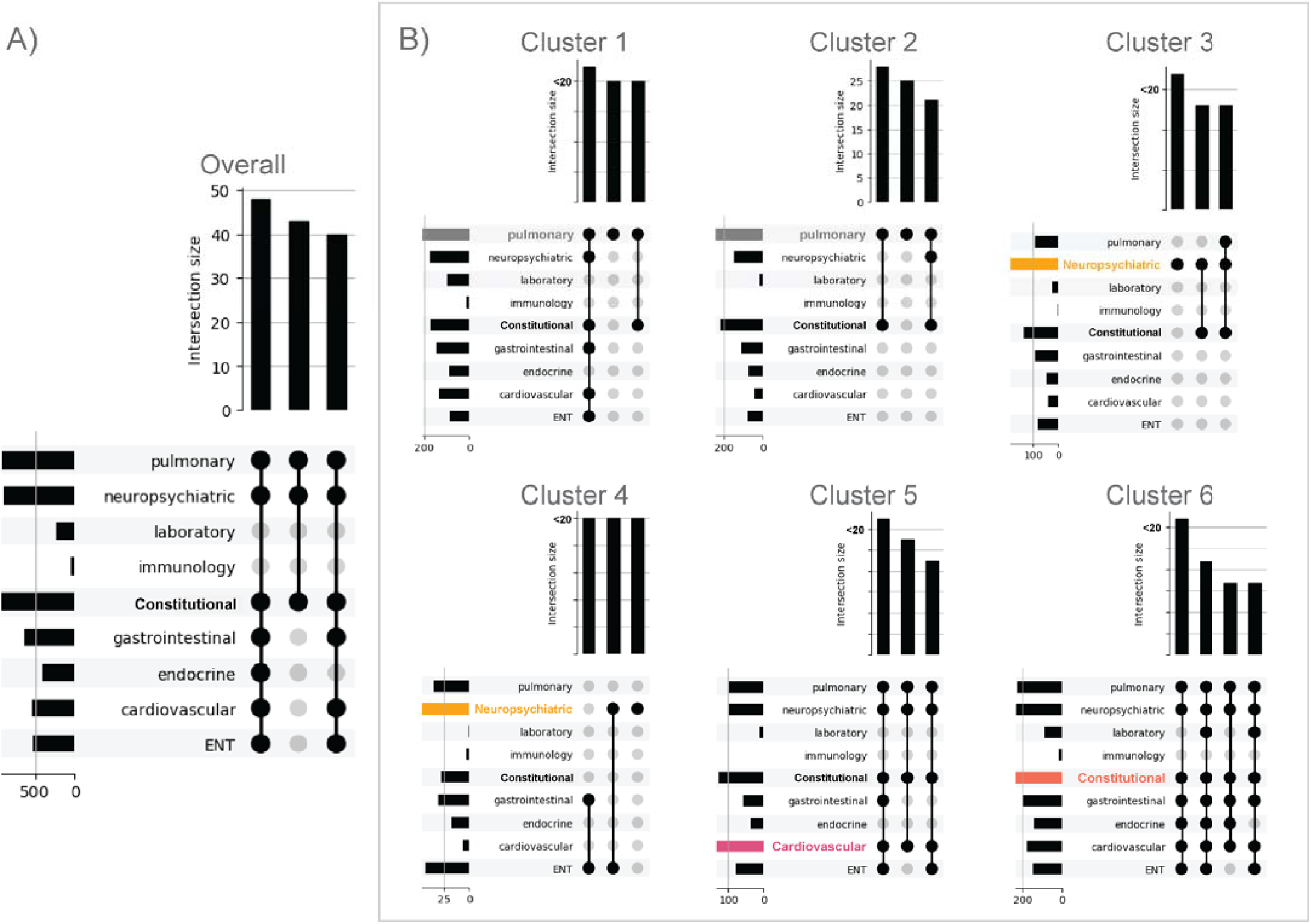
Phenotypically characterizing long COVID subtype clusters. Shown are the most frequently co-occurring high-level HPO categories for patients in the overall cohort (A) and for each of the 6 clusters (B). For the overall population of patients in data partner 1 and for each cluster, the frequency of each category of long COVID HPO terms (left) and the frequency of the three most common combinations of HPO categories (top) are shown. Notably, most clusters contain some widely shared features, but also distinguishing features such as symptoms in the pulmonary, neuropsychiatric, and cardiovascular systems. Data are shown as UpSet plots, which visualize set intersections in a matrix layout and show the counts of patients with the combination indicated by the black dots as bars above the matrix *(28)*. The most commonly occurring HPO category in each cluster is highlighted.

HPO terms were classified into these categories: cardiovascular, constitutional, endocrine, ear nose and throat (ENT), eye, gastrointestinal, immunology, laboratory, neuropsychiatric, pulmonary, and skin. The constitutional category encompasses symptoms and findings such as *Fatigue* (HP:0012378), *Night sweats* (HP:0030166), and *Xerostomia* (HP:0000217) that cannot be unambiguously assigned to a single organ system. UpSet plots *(28)* were used to visualize the salient characteristics of each cluster according to these categories. UpSet visualizations show not only the most common categories, but also the most common combinations of categories. For instance, in cluster 1, patients most commonly had HPO terms from the categories pulmonary, neuropsychiatric, general, gastrointestinal, cardiovascular, and ear nose throat (ENT), and the single most common category overall was pulmonary. Although there was some overlap in the distribution of features, the profiles of terms and categories were distinct for the six clusters (Figure 4).

### The six PASC clusters differ with respect to frequencies of clinical manifestations

Marked differences among groups were seen in the frequency with which certain symptoms were observed. For example, *Nasal Congestion* (HP:0001742) was frequent (∼31%) in cluster 4, and *Cough* (HP:0012735) was especially common (>60% of patients) in clusters 2 and 6 compared with the other clusters, although appreciable rates of *Cough* (HP:0012735) were seen among all clusters. Cardiac or potential cardiac signs and symptoms, such as *Palpitations* (HP:0001962), *Tachycardia* (HP:0001649), or *Chest pain* (HP:0100749), were relatively common in clusters 5 and 6 compared with the other clusters, although chest pain was also seen in ∼31% of cluster 2 patients. *Hypotension* (HP:0002615) was most common in cluster 6. *Pain* (HP:0012531) and *Fatigue* (HP:0012378) were relatively frequent in clusters 2, 3, and, particularly, clusters 5 and 6 (rates for these symptoms ranged from ∼56-79% in the latter two clusters). Cluster 6 was also notable for a high frequency of other constitutional symptoms, including *Fever* (HP:0001945), *Asthenia* (HP:0025406), and *Myalgia* (HP:0003326), as well as a number of gastrointestinal symptoms, such as *Abdominal pain* (HP:0002027), *Diarrhea* (HP:0002014), and *Nausea* (HP:0002018). *Vertigo* (HP:0002321) was common in cluster 5 (∼34%) and cluster 6 (∼25%). *Depression* (HP:0000716) and *Headache* (HP:0002315) were more common in clusters 3 and 6 versus other cohorts, and *Insomnia* (HP:0100785) was most frequent in cluster 6 (Figure 5).

**Figure 5.**
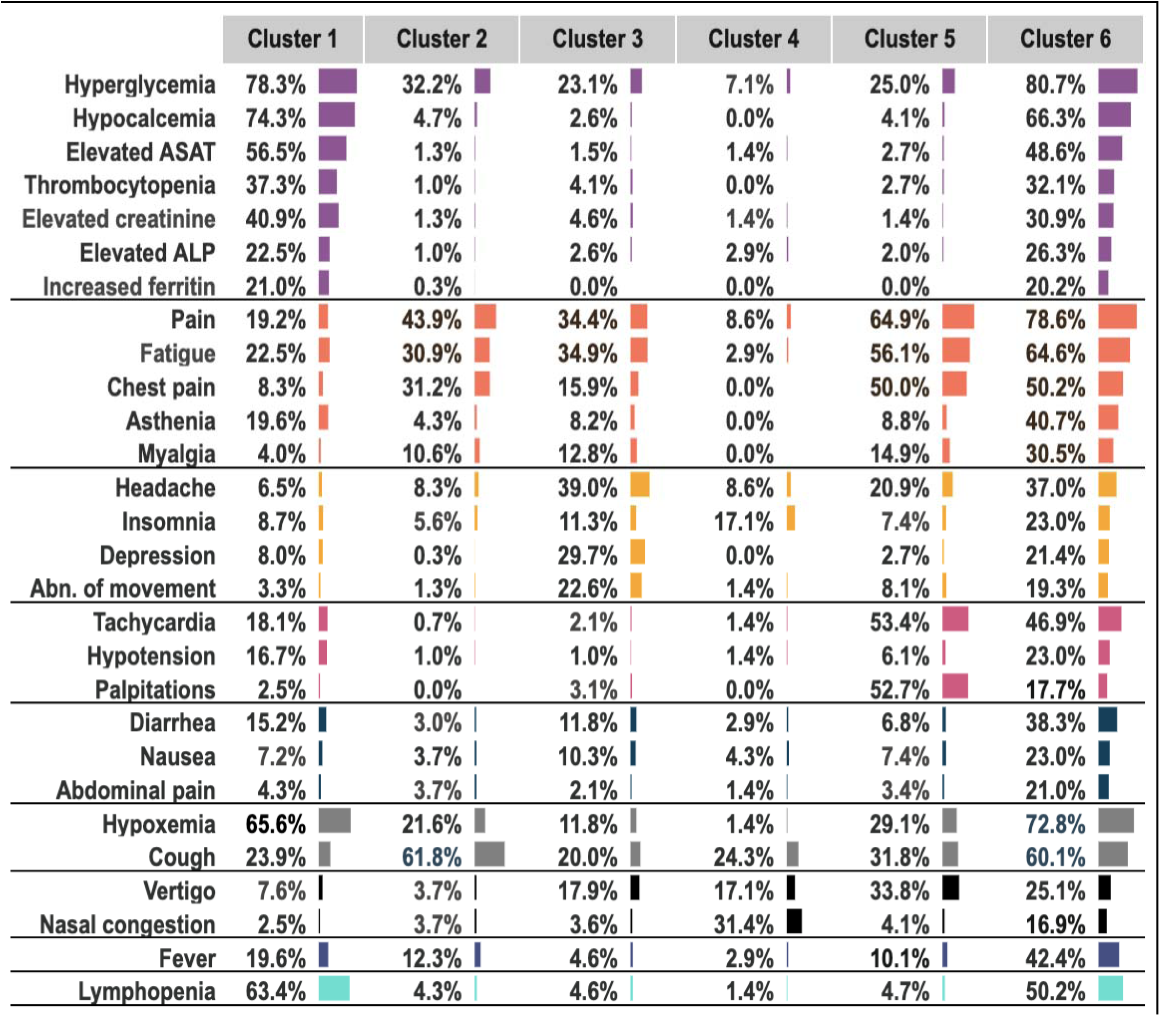
Summary of phenotypic feature distribution in the six clusters. The HPO terms corresponding to different phenotypic features are grouped in HPO categories shown on the left in this order: laboratory, constitutional, neuropsychiatric, cardiovascular, gastrointestinal, pulmonary, ENT, endocrine/metabolism, and immunological. Laboratory abnormalities are grouped together because of their association with severe COVID-19 (see text). HPO terms are shown if at least 20% of patients in at least one cluster had the corresponding phenotypic feature and if Pearson’s chi-squared test found a significant difference (p<0.00001) in the phenotypic feature distribution.

### Cluster 1 and 6 are characterized by manifestations suggesting increased clinical severity

Both advanced age and female sex have been associated with an increased risk of developing long COVID *(29)*. Interestingly, the average age in clusters 1 and 6 was higher than that in the other clusters, but the proportion of women in these clusters was lower than in three of the other four clusters. More patients in clusters 1 and 6 were in-patients during the bout of acute COVID-19 than patients in other clusters. Both clusters 1 and 6 showed a high frequency of post-acute COVID-19 laboratory abnormalities that have been associated with severe course of acute COVID-19, namely, *Lymphopenia* (HP:0001888), *Elevated circulating alanine aminotransferase concentration* (HP:0031964), *Increased circulating ferritin concentration* (HP:0003281), *Elevated circulating alkaline phosphatase concentration* (HP:0003155), *Hypocalcemia* (HP:0002901), and *Thrombocytopenia* (HP:0001873) *(30–35)*. This, and the fact that the average age was higher and the overall frequency of annotations with HPO terms was higher in these clusters (Supplemental Fig 1), suggests that clusters 1 and 6 may represent patients with residual manifestations of more severe COVID-19 and/or long COVID manifestations, although severity cannot unambiguously be inferred from EHR data.

### The six PASC clusters differ with respect to pre-existing comorbidities

To investigate how clinical features before or during COVID-19 infection correlated with cluster membership, we assessed the distribution across the six clusters of 44 clinical features determined prior to acute COVID-19 or during acute COVID-19. Of these, 19 displayed a statistically significant difference between clusters and are shown in Tables 2 and 3. Among parameters that were present before acute COVID-19 (Table 2), 13 differed significantly between clusters. Chronic lung disease, peripheral vascular disease, kidney disease, diabetes, coronary artery disease, heart failure, and acute kidney injury (AKI) were all more frequent in clusters 1 and 6 (Table 2). The risk of long COVID has been shown to be associated with the number of comorbidities *(36)*. Additionally, obesity, which has been shown to be a risk factor for long COVID *(37)*, was also more common in clusters 1 and 6. These observations are consistent with the notion that clusters 1 and 6 are composed of patients with more severe clinical manifestations, and that there may be different risk factors for clusters 2-5. Covariates during acute COVID-19 whose frequencies were higher in clusters 1 and 6 included acute kidney injury (AKI) and medications such as corticosteroids, remdesivir, and vasopressors that may be proxies for a severe clinical course (Table 3). Severity of acute COVID has been associated with risk of persistent symptoms in some studies *(38)*.

**Table 2.** Clinical features of patients before acute COVID-19 infection by cluster. The 13 of 35 clinical features present before COVID-19 infection (Supplemental Table S12) that were significantly overrepresented in clusters (chi squared p < 0.001 after Bonferonni correction) and the percent of patients in each cluster with each clinical feature are shown.

| Pre-existing Clinical Feature | Cluster 1 | Cluster 2 | Cluster 3 | Cluster 4 | Cluster 5 | Cluster 6 |
| --- | --- | --- | --- | --- | --- | --- |
| chronic lung disease | 37.2% | 20.0% | 21.6% | 20.3% | 16.3% | 37.4% |
| peripheral vascular disease | 7.3% | 1.0% | 1.5% | 1.4% | 3.4% | 11.1% |
| systemic corticosteroids | 61.3% | 49.3% | 48.5% | 37.7% | 41.5% | 71.6% |
| kidney disease | 27.0% | 3.0% | 4.6% | 4.3% | 2.7% | 22.6% |
| obesity | 58.8% | 44.3% | 48.5% | 39.1% | 37.4% | 66.3% |
| diabetes (uncomplicated) | 29.9% | 12.0% | 8.8% | 7.2% | 4.8% | 28.8% |
| coronary artery disease | 15.0% | 2.3% | 4.1% | 1.4% | 5.4% | 11.9% |
| diabetes (complicated) | 23.7% | 4.3% | 6.2% | 5.8% | 2.0% | 23.0% |
| hypertension | 46.7% | 25.0% | 28.9% | 21.7% | 17.0% | 49.8% |
| congestive heart failure | 8.8% | 2.0% | 1.0% | 0.0% | 0.7% | 7.8% |
| heart failure | 11.7% | 2.0% | 1.5% | 1.4% | 2.0% | 10.3% |
| depression | 16.4% | 16.0% | 35.6% | 15.9% | 15.0% | 29.2% |
| AKI | 22.6% | 0.7% | 2.6% | 1.4% | 0.7% | 14.0% |

**Table 3.**
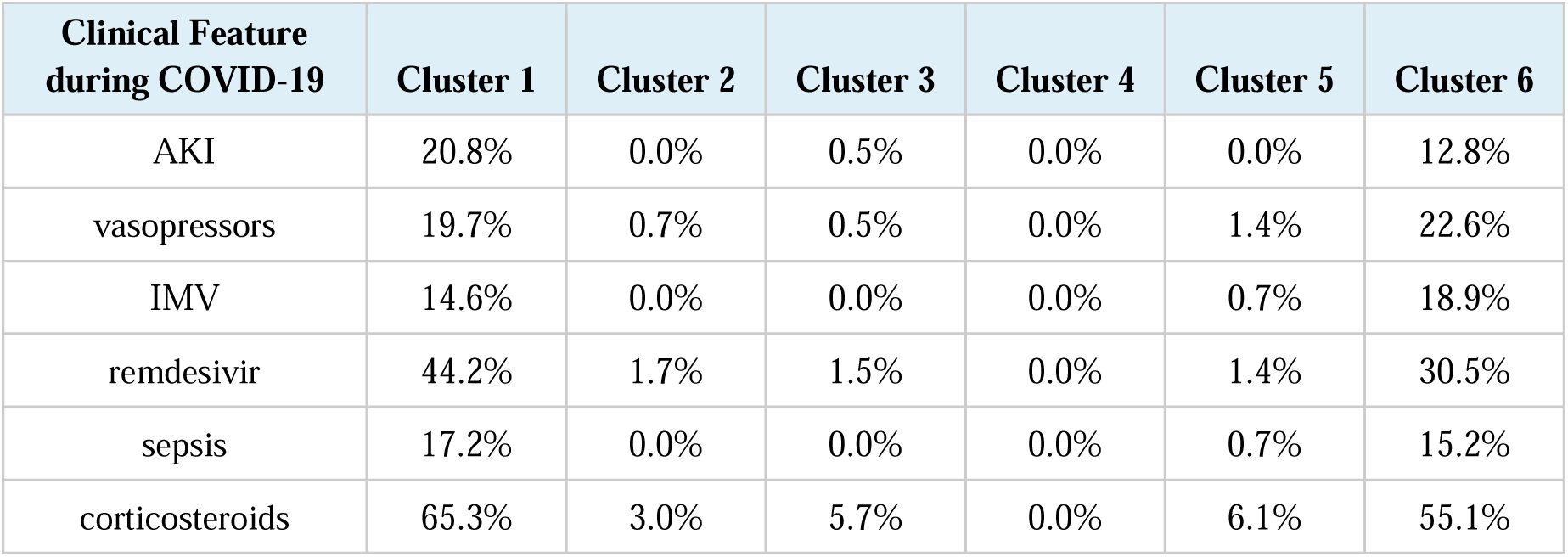
Clinical features of patients during acute COVID-19 infection by cluster. The 6 of 9 clinical features present during COVID-19 infection (Supplemental Table S13) that were significantly overrepresented in clusters (chi squared p < 0.001 after Bonferonni correction) and the percent of patients in each cluster with each clinical feature are shown.

### Generalizability of clusters to new data partners

The results presented in the previous sections were generated with data from data partner 1. We assessed the generalizability of the clustering results for four additional data partners (data partners 2-5, Figure 1) by comparing each patient from these data partners to the patients in each cluster from data partner 1 and also to randomly permuted clusters (Methods). If the clusters in data partner 1 did not generalize at all to other data partners, we would expect that patients from other data partners would be equally similar to the patients of any of the clusters in data partner 1.

We observed that patients from data partners 2-5 were much more similar to clusters from data partner 1 compared to randomly permuted clusters. The mean similarity ranged from 0.179 to 0.182 for test data partners 2-5 for the randomly permuted clusters, but the observed mean similarities to the original clusters at data partner 1 ranged from 0.270 to 0.300, corresponding to z-scores of 150 to 266. The mean similarity score for the randomly permuted clusters was never as high as the observed score over 1000 permutations, corresponding to an empirical p-value of less than 0.001 for each of the data partners 2-5. This strongly suggests that clusters identified in data partner 1 generalize to patients from other data partners (Table 4).

**Table 4.** Generalizability of clusters in patients from new data partners. The similarity of patients from test data partners 2-5 to patients from data partner 1’s clusters and to patients from randomly permuted clusters was measured as in Fig 2. For each test data partner, the average similarity of its patients to the best matching randomly permuted cluster and to the best matching cluster from data partner 1 are shown along with the Z-score and p-value. The empirical p-value reflects the number of times that the similarity of a permuted dataset was higher than that of the observed clusters (this never occurred).

| Test data partner | Similarity to permuted clusters | Observed mean similarity | Z-score | Empirical p-value |
| --- | --- | --- | --- | --- |
| 2 | $0.179 \pm 0.000351$ | 0.270 | 261.0 | $< 0.001$ |
| 3 | $0.179 \pm 0.000387$ | 0.271 | 236.3 | $< 0.001$ |
| 4 | $0.180 \pm 0.000355$ | 0.274 | 266.0 | $< 0.001$ |
| 5 | $0.182 \pm 0.000787$ | 0.300 | 149.7 | $< 0.001$ |

**Table 5.** Recorded deaths according to cluster. Data partner 1 was the source of data for generating the six clusters. Patients from data partners 2-5 were assigned to these clusters (Methods). Deaths recorded in the EHR and total number of patients are shown.

| Cluster | Data partner 1 |  | Data partners 2-5 |  |
| --- | --- | --- | --- | --- |
|  | deaths | total | deaths | total |
| 1 | 20 | 276 | 19 | 215 |
| 2 | 1 | 301 | 0 | 37 |
| 3 | 0 | 195 | 0 | 128 |
| 4 | 0 | 70 | 0 | 50 |
| 5 | 0 | 148 | 0 | 167 |
| 6 | 14 | 243 | 13 | 634 |

### Clusters 1 and 6 are characterized by higher mortality reproducibly across data partners 1-5

Because of the indications that clusters 1 and 6 may be characterized by greater clinical severity, we assessed recorded mortality in the time period subsequent to acute COVID-19. In data partner 1, all deaths except 1 were recorded in patients assigned to either cluster 1 or 6 (97%). We assigned patients from data partners 2-5 to the original six clusters according to the maximum mean similarity of patients in those clusters (Methods). In these patients, all cases of recorded mortality occurred in patients assigned to clusters 1 and 6. We performed a chi-squared test of the null hypothesis that the proportion of mortalities in the clusters was uniform. The observed correlation between mortality and cluster membership was statistically significant for the analysis of clustered patients in data partner 1 (□ = *5 × 10^−5^*) and in data partners 2-5 (□ = *5 × 10^−5^*) using a Fisher’s exact test calculated by the Monte Carlo method.

## DISCUSSION

According to the World Health Organization, approximately 10-20% of patients with COVID-19 may experience new-onset, lingering or recurrent clinical symptoms after acute infection. This has been termed ‘post-acute sequelae of SARS-CoV-2 infection’ (PASC) or long COVID. Definitions of long COVID in the literature vary, and the frequencies and time course of phenotypic manifestations following acute COVID-19 are highly heterogeneous.*(19)* This observation raises the question of whether long COVID can be stratified into well delineated and reproducible subtypes, or whether the degree of heterogeneity is so high that stratification is impossible. This is critically relevant for defining sub-cohorts in clinical research studies such as the NIH program “Researching COVID to Enhance Recovery (RECOVER),” and for identifying candidate therapeutics. ML clustering methods offer a data-driven approach to stratification of patients that can reveal such subtypes in the face of this new and heterogeneous disease.

Evidence available prior to our study suggests that important clinical differences do exist that influence the susceptibility to subsequent complications of COVID-19. For instance, although males are more likely to be hospitalized or die with acute COVID-19, females are more likely to develop long COVID *(39)*. It is possible that the pathophysiology of long COVID may be multifactorial in origin. Conceivably, the biological underpinnings of long COVID may vary among individuals as a function of baseline risk factors, resulting in different general phenotypes of long COVID, the treatment or prevention of which may need to be specifically tailored using precision medicine in order to achieve optimal outcomes. As a first step, we sought to use unsupervised learning to delineate potential subtypes of patients with long COVID with differing clinical characteristics. We identified six published studies that present clusters from either patient-reported data (in four studies) or manually recorded clinical data (two studies) with cohorts of between 145 and 3762 patients. The studies report two or three clusters based on different types of input data, making study comparison challenging. None of the studies were based on EHR data and no assessment of generalizability to other data partners was presented *(18, 29, 40–43)*.

Here we have presented a novel method for semantic clustering of long COVID patients based on HPO-encoded EHR data. We further present a method for assessing generalizability of the identified subtypes or clusters across different data contributing sites. Ontology-based algorithms differ from machine learning and other algorithms in many ways. Coding numerical data with HPO implies that parameters are simplified into categories. Although this loss of numerical data reduces precision in data granularity, simplification allows powerful simultaneous analysis of all phenotypic observations using semantic similarity that can take the relatedness of concepts into account.

Our method for assessing patient-patient similarity using the Phenomizer algorithm generates an essentially continuous similarity value from arbitrary sets of HPO terms that characterize any two patients. An alternative method would be to encode the 287 HPO terms as a 287-dimensional feature vector and to measure similarity for example using dot product (cosine) of these vectors. The Phenomizer algorithm has several advantages over the feature vector method: it does not suffer from sparse count issues that may make clustering less robust *(44)*, and it takes advantage of the similarity between individual items using the structure of the HPO in a way that a feature vector cannot *(26)*. This approach has proven powerful both in the support of differential diagnosis of rare disease and in efforts to enable longitudinal analysis of EHR data as a means of identifying gene-phenotype associations with Mendelian forms of epilepsy *(45, 46)*, but has never before been applied in the context of infectious disease EHR data and methods for assessing generalizability have not previously been presented.

We have shown that unsupervised learning based on semantic clustering identifies phenotypic profiles that are reproducible across data partners with a high degree of statistical significance. The six clusters that emerged demonstrated non-uniform frequencies of symptoms and clinical findings across an array of features, spanning constitutional/systemic symptoms and pain, cardiac, respiratory, gastrointestinal, and neurologic symptom domains, with some degree of overlap but clear distinctions between various groups. We interpret our clusters 1 and 6 as comprising patients with a severe course of acute COVID-19 because of the higher hospitalization rates (Table 1) and the higher rates of mechanical ventilation and use of medication such as vasopressors that indicate a relatively severe course (Table 3). It is possible that these clusters represent a subtype of long COVID that results from severe acute COVID-19. Interestingly, cluster 1 was male-predominant (59.1%) and cluster 6 was female predominant (58.0%). The higher rates of mortality and most pre-existing comorbidities in patients from clusters 1 and 6 are in accordance with the notion of more severe clinical courses. Our results show that these subgroups tended to be affected by a wider range of clinical complications in the post-acute course, because, for instance, the most common profile of HPO terms involved six of nine clinical categories in cluster 1 and seven of nine in cluster 6 (Figure 4). Our findings confirm and extend previous findings of a steeper risk gradient for long COVID manifestations that increases according to the severity of the acute COVID-19 infection *(47)*.

The relatively high rate of pre-COVID corticosteroid use in our study (with the lowest rate being 37.7% in cluster 4 and the two highest rates 61.3% in cluster 1 and 71.6% in cluster 6) is striking. Dexamethasone use was associated with lower 28-day mortality among those who were receiving either invasive mechanical ventilation or oxygen but not among those receiving no respiratory support *(48)*. However, methylprednisolone use may be associated with increased mortality and more severe neuromuscular weakness in some patients with acute respiratory syndrome (ARDS) *(49)* and there are reasons to believe that protracted corticosteroid therapy could contribute to the development of some long COVID manifestations such as fatigue, myopathy, neuromuscular weakness, and psychiatric symptoms *(50)*. However, future work will be needed to determine what causal role, if any, steroid use has in the development of long COVID.

A substantial body of evidence documents a sex difference in the severity of acute COVID-19, with a more favorable course of the disease in women compared to men regardless of age *(51)*. Emerging evidence suggests that the clinical manifestations of long COVID may also be characterized by sex differences *(52–54)*. Our results show a cluster with predominantly hospitalized and male patients (cluster 1) and other clusters with predominantly non-hospitalized and female patients (clusters 3 and 4), which suggests that males and females may differ with respect to long COVID manifestations. A focused, prospective study could help to clarify the extent potential sex differences in long COVID.

We suggest that analogous algorithms could be used to evaluate data gathered from prospective studies of long COVID patients to extend and deepen our characterization of phenotypic clusters by including data that are currently difficult to ascertain reliably from EHR data, including symptoms such as *Asthenia* (HP:0025406) or *Exertional dyspnea* (HP:0002875) and radiology findings (which are typically not represented using structured fields in EHR data and are underrepresented in OMOP datasets). The recently released Phenopacket Schema of the Global Alliance for Genomics and Health (GA4GH) provides a standardized way to record clinical findings including phenotypic features, measurements, biospecimens, and medical actions over the time course of a disease as a computational case report *(55)*. Recording clinical data with the Phenopacket Schema would promote data sharing and comparability of results from different studies.

### Study limitations

While our study provides insight into the variability and natural history of long COVID, there are limitations that should be considered. While the U09.9 code provides a simple inclusion criterion, its application in health systems across the country is not uniform and may differ from one data partner to another. Also, since the use of the code began only recently, patients with long COVID that were diagnosed prior to the introduction of the code are not included, limiting our ability to compare the current clinical manifestations with those observed earlier in the pandemic before widespread vaccination and with different distributions of SARS-CoV2 strains and variants. However, in a pilot study in Denmark, coding with U09.9 was found to have a positive predictive value of 94% for long COVID *(56)*.

Our ability to capture clinical manifestations of long COVID is limited by the accessibility of clinical data in EHR systems. Of the 287 HPO terms we identified as being used in published cohort studies on long COVID,*(19)* only 116 were identified in our data. The reasons for this presumably include unstructured data such as symptoms and radiological findings that are not well represented in the OMOP data that is the source of our data. Examples include *Gaze-evoked nystagmus* (HP:0000640), *Pericardial effusion* (HP:0001698), and *Exercise intolerance* (HP:0003546) that are typically diagnosed using specialist examinations or medical history that may not be easily coded in structured EHR fields. Additionally, several common manifestations of long COVID, including dysautonomia *(57)*, are less documented in EHR data in part due to the difficulties in recognizing these illnesses clinically and the fact that relevant findings may not be well represented in structured fields including the OMOP data available in N3C.

Our study uses the newly minted ICD code U09.9 to identify patients with PASC/long COVID. At the time of this writing, a relatively small number of affected patients was available for analysis. Furthermore, the population defined by these patients is not fully representative of the American population; for instance, the proportion of African Americans in our study (∼5%) is lower than the proportion of African Americans among the entire population. As more data accrues, future work will be required to characterize the role of social determinants of health that are confounded with race in our society in determining long COVID subtypes. It is likely that many additional long COVID patients are present in the N3C dataset who have not received the U09.9 diagnosis code, and it is possible that this fact could introduce a bias into the data analyzed in this study. Additionally, the group of patients who present for medical care for long COVID symptoms and receive a U09.9 diagnostic code may not be representative of the entire population of patients with long COVID manifestations.

Our exploration of k-means clustering results with different values of k from 2 to 8 showed that increasing the number of clusters tended to subdivide existing clusters hierarchically. Although numerous methods for determining the ‘best’ number of clusters are available, there is no objective definition of optimum that applies to all applications, and the choice of k is perforce subjective in nature. Our main findings of generalizable phenotypic clusters pertain also for values of k of 4 and 5 (Supplemental Figure S2-S3).

## Conclusions

We have presented a novel algorithm for semantic clustering that identifies patient similarity by transforming EHR data to phenotypic profiles using the HPO, and reviewed long COVID subtypes that show a statistically significant degree of generalizability of clusters across different medical centers. There was a significant association of cluster membership with a range of pre-existing conditions and with measures of severity during acute COVID-19. Two of the clusters were associated with severe manifestations and displayed increased mortality. Additionally, we show that the identified clusters were generalizable across different hospital systems and that the increased mortality rate was consistently observed in two of the clusters. Semantic phenotypic clustering could provide a basis for assigning patients to stratified subgroups for natural history or therapy studies.

## MATERIALS AND METHODS

The N3C data transfer to NCATS is performed under a Johns Hopkins University Reliance Protocol #IRB00249128 or individual site agreements with NIH. The N3C Data Enclave is managed under the authority of the NIH; information can be found at https://ncats.nih.gov/n3c/resources.

### Setting

We obtained patient data from the National COVID Cohort Collaborative (N3C; covid.cd2h.org). N3C aggregates and harmonizes EHR data across multiple clinical organizations in the United States, including the Clinical and Translational Science Awards (CTSA) Program hubs. N3C harmonizes EHR data across four clinical data models and provides a unified analytical platform in which data are encoded using the Observational Medical Outcomes Partnership (OMOP)*(27)* version 5.3.1.

### Cohort

The Centers for Disease Control (CDC) announced an International Classification of Diseases, version 10 (ICD-10) code (U09.9) for emergency/provisional use on June 30, 2021. The code represents Post COVID-19 condition, unspecified. Use of the code was approved for implementation effective October 1, 2021. The code should be used for patients with a history of probable or confirmed SARS CoV-2 infection who are identified with a post-COVID condition. The data freeze date was March 16, 2022. Only patients with an initial COVID-19 diagnosis within the Enclave were included in the cohort. At the time of the data freeze for this analysis, 21 participating data partners were using the code, and a total of 5645 patients were coded in this way.

### Human Phenotype Ontology (HPO)

The HPO is a rich representation of the diversity of phenotypic features associated with human disease and is the de facto standard for the computational analysis and exchange of phenotype data in human genetics *(20, 58–62)*. The HPO comprises over 16,000 terms that denote specific phenotypic abnormalities at increasingly specific granularity, for example, *Atrial septal defect* (HP:0001631) and *Interrupted inferior vena cava with azygous continuation* (HP:0011671). We recently identified 287 unique clinical findings reported in cohorts of patients with long COVID and mapped them to existing HPO terms and in some cases created new HPO terms to cover COVID-specific features such as *Pseudo-chilblains on toes* (HP:0034036) *(19)*. The 2020-08-11 release of the HPO was used in our study.

### Mapping OMOP codes to HPO terms

To obtain mappings between standard OMOP condition concept identifiers and HPO concepts, we used OMOP2OBO (https://github.com/callahantiff/OMOP2OBO) and LOINC2HPO *(63, 64)*. The OMOP2OBO algorithm creates and validates mappings between OMOP terminology concepts and concepts from the Open Biomedical Ontologies *(65)*, using a variety of alignment strategies and with varying levels of confidence. For this project, we filtered the v1.0.0 release of mappings to only include exact 1:1 mappings at the concept level. This mapping set aligned 4,767 OMOP concept IDs to 3,804 unique HPO concepts (1.25 OMOP concept IDs/HPO concept). To apply LOINC2HPO mappings from OMOP to HPO concepts, we reimplemented the LOINC to HPO mappings in the N3C Enclave. For any HPO term that was among the 287 HPO terms associated with long COVID, we determined for each patient in our study group the LOINC codes present in the measurement OMOP table determined to be ‘low’, ‘high’, or ‘positive’ compared to the reference range for the test in question, and assigned the HPO term to the patient if the test occurred during the long COVID period for that patient (starting 21 days after diagnosis of acute COVID-19 for outpatients, and 21 days after hospitalization for inpatients).

### Specificity-weighted fuzzy phenotype matching

We previously developed a method called Phenomizer for clinical diagnostics that uses the semantic structure of the HPO to weight clinical features on the basis of specificity and to identify those clinical features that best distinguish among the top candidate differential diagnoses *(26)*. The algorithm represents the clinical specificity of a finding as the information content (IC) of a term. Given a set of diseases of interest in the differential diagnosis process, the frequency of each HPO term is defined as the proportion of diseases in a database that are annotated by the term or any of its descendent terms (for instance, the HPO resource currently comprises 8,260 Mendelian diseases) *(21)*. The IC is then defined as the negative natural logarithm of the term frequency *(66)*. The true path rule applies to all terms in the HPO. That is, if a disease is annotated to the term □, it is implicitly annotated to all ancestors of □ recursively (for instance, Marfan syndrome is annotated to *Aortic root aneurysm* (HP:0002616), and it is therefore implicitly annotated to the parent term *Thoracic aortic aneurysm* (HP:0012727) and its parent term *Aortic aneurysm* (HP:0004942), and so on. Thus, the IC of terms increases as we move from the root term of the HPO ontology to the more specific descendent terms.

To define the similarity between any two HPO terms □_*1*_ and □_*2*_, we find the most specific common ancestor of □_*1*_ and □_*2*_ (which we call the Most Informative Common Ancestor of □_*1*_ and □_*2*_, MICA(*t*_l_, t_2_) in the hierarchy and calculate its IC as IC(MICA(t_l_,*t*_2_). In essence, this procedure leverages the ontological structure of the HPO to perform specificity-weighted fuzzy matching.

In the Phenomizer algorithm, the similarity between a set of query terms (symptoms, signs, etc.) entered by a physician for an individual case is used to calculate a similarity score for each of the diseases in the HPO database as an aid in differential diagnosis. In the current work, we adapt this algorithm to implement semantic phenotypic-based clustering by using the Phenomizer framework to calculate a matrix of pairwise phenotypic similarities between all patients in the long COVID cohort. In the following, we represent the set of □ long COVID patients as *p*_l_, *p*_2_,…,*p*_*n*_ ∈ *P*. The set of □ HPO terms associated with patient □ is represented as *t*_l_,…,*t*_*m*_ ∈ *p*_*i*_. Then the similarity from patient □_□_to □_□_is calculated as

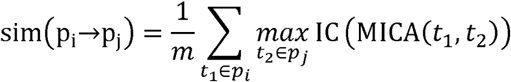

This equation is not symmetric, so the final similarity score is calculated as

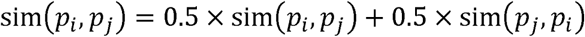

### k-means clustering

For □ patients, we calculated a similarity matrix X^*n*×*n*^ using the Phenomizer algorithm. We then applied k-means clustering to partition the patients into □ clusters, denoted *c*_l_,*c*_2_, …,*c*_*c*_, where □_□_is the set of □_□_objects in cluster □ and □ is the number of clusters (a user-chosen hyperparameter). Using a previously described method, □ cluster centroids were chosen such that centroids were distant from one another *(67)*. Clusters were then formed iteratively such that the Euclidean distance between the vector that represents any object and the centroid vector of its cluster was at least as small as that between the object and any of the other clusters. In each iteration, objects were moved to the cluster with the closest centroid, following which the centroids were recalculated until no further improvement was obtained or the maximum number of 100 iterations was reached *(68)*.

The k-means clustering method does not determine the ‘optimal’ number of clusters. We used the elbow method to choose the number of clusters. This method computes the total within-cluster sum of squares error (SSE) for each candidate number of clusters. The SSE is plotted against the number of clusters and an ‘elbow’ in the curve is used to determine the number of clusters.

### Assessing cluster reproducibility between data partners

We first performed clustering on patients from the data partner with the greatest number of U09.9 long COVID patients. For brevity, we will refer to this as data partner 1. We then assessed reproducibility of clustering results in data partners 2-5 as explained below. This approach was chosen given the inherent challenge owing to the lack of a generally applicable method for assessing any given clustering approach *(69–71)*. For brevity, we will refer to these data partners 2-5 as the test data partners.

The HPO terms for patients from data partner 1 and their assignment to k-means clusters were recorded. We reasoned that if the clustering results in data partner 1 are generalizable, then patients of the test data partners will tend to display more similarity to one or other cluster of data partner 1 than one would expect by chance. Assuming we have □ clusters from data partner 1, then a weighted similarity vector can be calculated for each patient □ from a test data partner as [*p*_1_, *p*_2_, …, *p*_*k*_] If the patient is equally similar to each of the □ clusters, then 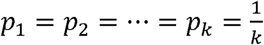. If, on the other hand, the patient is much more similar to one of the clusters, say cluster □, then we expect 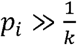. We therefore define the test statistic *p*_*max*_ = *max*_*i*_ *p*_*i*_ for patient □. To assess generalizability, we calculate □_□□□_ for each patient □ in the test data partner and take the mean value of □_□□□_over all patients in the test data partner as our test statistic 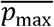. To generate a null distribution of this statistic, we create 1,000 permuted cluster assignments by assigning each patient from data partner 1 uniformly at random to one of the k clusters. We compute the test statistic for each of these random cluster assignments and record the mean and standard deviation of these values. We present the results as a z score calculated as 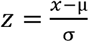.

### Assessing covariate distribution

The HPO terms assessed in the above procedures were derived from clinical data at least 21 days after the initial bout of COVID-19. We analyzed additional clinical covariates covering items such as comorbidities and medications prior to and during acute COVID-19 (Supplemental Tables S2-S3). Categorical variables were assessed with a chi-squared test if at least five counts were present for each cell of the contingency table and numerical variables were assessed with one-way ANOVA. Analysis was done using R version 3.5.1.

## Data Availability

Data are available by application to the N3C Data Enclave, which is managed under the authority of the NIH; information can be found at https://ncats.nih.gov/n3c/resources.

## Acknowledgments

This study is part of the NIH Researching COVID to Enhance Recovery (RECOVER) Initiative (https://recovercovid.org/), which seeks to understand, treat, and prevent the post-acute sequelae of SARS-CoV-2 infection (PASC) and; and was conducted under the N3C DUR RP-5677B5. The views and conclusions contained in this document are those of the authors and should not be interpreted as representing the official policies, either expressed or implied, of the NIH.

Medical Authorship was determined using ICMJE recommendations.

The analyses described in this publication were conducted with data or tools accessed through the NCATS N3C Data Enclave covid.cd2h.org/enclave and supported by NCATS U24 TR002306. This research was possible because of the patients whose information is included within the data from participating organizations (covid.cd2h.org/dtas) and the organizations and scientists (covid.cd2h.org/duas) who have contributed to the on-going development of this community resource.*(72)*

The N3C data transfer to NCATS is performed under a Johns Hopkins University Reliance Protocol # IRB00249128 or individual site agreements with NIH. The N3C Data Enclave is managed under the authority of the NIH; information can be found at https://ncats.nih.gov/n3c/resources.

We gratefully acknowledge the following core contributors to N3C: Anita Walden, Leonie Misquitta, Joni L. Rutter, Kenneth R. Gersing, Penny Wung Burgoon, Samuel Bozzette, Mariam Deacy, Christopher Dillon, Rebecca Erwin-Cohen, Nicole Garbarini, Valery Gordon, Michael G. Kurilla, Emily Carlson Marti, Sam G. Michael, Lili Portilla, Clare Schmitt, Meredith Temple-O’Connor, David A. Eichmann, Warren A. Kibbe, Hongfang Liu, Philip R.O. Payne, Emily R. Pfaff, Peter N. Robinson, Joel H. Saltz, Heidi Spratt, Justin Starren, Christine Suver, Adam B. Wilcox, Andrew E. Williams, Chunlei Wu, Davera Gabriel, Stephanie S. Hong, Kristin Kostka, Harold P. Lehmann, Michele Morris, Matvey B. Palchuk, Xiaohan Tanner Zhang, Richard L. Zhu, Benjamin Amor, Mark M. Bissell, Marshall Clark, Andrew T. Girvin, Stephanie S. Hong, Kristin Kostka, Adam M. Lee, Robert T. Miller, Michele Morris, Matvey B. Palchuk, Kellie M. Walters, Will Cooper, Patricia A. Francis, Rafael Fuentes, Alexis Graves, Julie A. McMurry, Shawn T. O’Neil, Usman Sheikh, Elizabeth Zampino, Katie Rebecca Bradwell, Andrew T. Girvin, Amin Manna, Nabeel Qureshi, Christine Suver, Julie A. McMurry, Carolyn Bramante, Jeremy Richard Harper, Wenndy Hernandez, Farrukh M Koraishy, Amit Saha, Satyanarayana Vedula, Johanna Loomba, Andrea Zhou, Steve Johnson, Evan French, Alfred (Jerrod) Anzalone, Umit Topaloglu, Amy Olex, Hythem Sidkey. Details of contributions available at covid.cd2h.org/acknowledgements

We acknowledge support from many grants; the content is solely the responsibility of the authors and does not necessarily represent the official views of the N3C Program, the NIH or other funders. In addition, access to N3C Data Enclave resources does not imply endorsement of the research project and/or results by NIH or NCATS.

We acknowledge the following institutions whose data is released or pending:

## Available

Advocate Health Care Network — UL1TR002389: The Institute for Translational Medicine (ITM) • Boston University Medical Campus UL1TR001430: Boston University Clinical and Translational Science Institute • Brown University — U54GM115677: Advance Clinical Translational Research (Advance-CTR) • Carilion Clinic — UL1TR003015: iTHRIV Integrated Translational health Research Institute of Virginia • Charleston Area Medical Center — U54GM104942: West Virginia Clinical and Translational Science Institute (WVCTSI) • Children’s Hospital Colorado — UL1TR002535: Colorado Clinical and Translational Sciences Institute • Columbia University Irving Medical Center — UL1TR001873: Irving Institute for Clinical and Translational Research • Duke University — UL1TR002553: Duke Clinical and Translational Science Institute • George Washington Children’s Research Institute — UL1TR001876: Clinical and Translational Science Institute at Children’s National (CTSA-CN) • George Washington University — UL1TR001876: Clinical and Translational Science Institute at Children’s National (CTSA-CN) • Indiana University School of Medicine — UL1TR002529: Indiana Clinical and Translational Science Institute • Johns Hopkins University — UL1TR003098: Johns Hopkins Institute for Clinical and Translational Research • Loyola Medicine — Loyola University Medical Center • Loyola University Medical Center — UL1TR002389: The Institute for Translational Medicine (ITM) • Maine Medical Center — U54GM115516: Northern New England Clinical & Translational Research (NNE-CTR) Network • Massachusetts General Brigham — UL1TR002541: Harvard Catalyst • Mayo Clinic Rochester — UL1TR002377: Mayo Clinic Center for Clinical and Translational Science (CCaTS) • Medical University of South Carolina — UL1TR001450: South Carolina Clinical & Translational Research Institute (SCTR) • Montefiore Medical Center — UL1TR002556: Institute for Clinical and Translational Research at Einstein and Montefiore • Nemours — U54GM104941: Delaware CTR ACCEL Program • NorthShore University HealthSystem — UL1TR002389: The Institute for Translational Medicine (ITM) • Northwestern University at Chicago — UL1TR001422: Northwestern University Clinical and Translational Science Institute (NUCATS) • OCHIN — INV-018455: Bill and Melinda Gates Foundation grant to Sage Bionetworks • Oregon Health & Science University — UL1TR002369: Oregon Clinical and Translational Research Institute • Penn State Health Milton S. Hershey Medical Center — UL1TR002014: Penn State Clinical and Translational Science Institute • Rush University Medical Center — UL1TR002389: The Institute for Translational Medicine (ITM) • Rutgers, The State University of New Jersey — UL1TR003017: New Jersey Alliance for Clinical and Translational Science • Stony Brook University — U24TR002306 • The Ohio State University UL1TR002733: Center for Clinical and Translational Science • The State University of New York at Buffalo — UL1TR001412: Clinical and Translational Science Institute • The University of Chicago — UL1TR002389: The Institute for Translational Medicine (ITM) • The University of Iowa — UL1TR002537: Institute for Clinical and Translational Science • The University of Miami Leonard M. Miller School of Medicine — UL1TR002736: University of Miami Clinical and Translational Science Institute • The University of Michigan at Ann Arbor — UL1TR002240: Michigan Institute for Clinical and Health Research • The University of Texas Health Science Center at Houston — UL1TR003167: Center for Clinical and Translational Sciences (CCTS) • The University of Texas Medical Branch at Galveston — UL1TR001439: The Institute for Translational Sciences • The University of Utah — UL1TR002538: Uhealth Center for Clinical and Translational Science • Tufts Medical Center — UL1TR002544: Tufts Clinical and Translational Science Institute • Tulane University — UL1TR003096: Center for Clinical and Translational Science • University Medical Center New Orleans — U54GM104940: Louisiana Clinical and Translational Science (LA CaTS) Center • University of Alabama at Birmingham — UL1TR003096: Center for Clinical and Translational Science • University of Arkansas for Medical Sciences — UL1TR003107: UAMS Translational Research Institute • University of Cincinnati — UL1TR001425: Center for Clinical and Translational Science and Training • University of Colorado Denver, Anschutz Medical Campus — UL1TR002535: Colorado Clinical and Translational Sciences Institute • University of Illinois at Chicago — UL1TR002003: UIC Center for Clinical and Translational Science • University of Kansas Medical Center — UL1TR002366: Frontiers: University of Kansas Clinical and Translational Science Institute • University of Kentucky — UL1TR001998: UK Center for Clinical and Translational Science • University of Massachusetts Medical School Worcester — UL1TR001453: The UMass Center for Clinical and Translational Science (UMCCTS) • University of Minnesota — UL1TR002494: Clinical and Translational Science Institute • University of Mississippi Medical Center — U54GM115428: Mississippi Center for Clinical and Translational Research (CCTR) • University of Nebraska Medical Center — U54GM115458: Great Plains IDeA-Clinical & Translational Research • University of North Carolina at Chapel Hill — UL1TR002489: North Carolina Translational and Clinical Science Institute • University of Oklahoma Health Sciences Center — U54GM104938: Oklahoma Clinical and Translational Science Institute (OCTSI) • University of Rochester — UL1TR002001: UR Clinical & Translational Science Institute • University of Southern California — UL1TR001855: The Southern California Clinical and Translational Science Institute (SC CTSI) • University of Vermont — U54GM115516: Northern New England Clinical & Translational Research (NNE-CTR) Network • University of Virginia UL1TR003015: iTHRIV Integrated Translational health Research Institute of Virginia • University of Washington — UL1TR002319: Institute of Translational Health Sciences • University of Wisconsin-Madison — UL1TR002373: UW Institute for Clinical and Translational Research • Vanderbilt University Medical Center — UL1TR002243: Vanderbilt Institute for Clinical and Translational Research • Virginia Commonwealth University — UL1TR002649: C. Kenneth and Dianne Wright Center for Clinical and Translational Research • Wake Forest University Health Sciences — UL1TR001420: Wake Forest Clinical and Translational Science Institute • Washington University in St. Louis — UL1TR002345: Institute of Clinical and Translational Sciences • Weill Medical College of Cornell University — UL1TR002384: Weill Cornell Medicine Clinical and Translational Science Center • West Virginia University — U54GM104942: West Virginia Clinical and Translational Science Institute (WVCTSI)

## Submitte

Icahn School of Medicine at Mount Sinai — UL1TR001433: ConduITS Institute for Translational Sciences • The University of Texas Health Science Center at Tyler — UL1TR003167: Center for Clinical and Translational Sciences (CCTS) • University of California, Davis — UL1TR001860: UCDavis Health Clinical and Translational Science Center • University of California, Irvine — UL1TR001414: The UC Irvine Institute for Clinical and Translational Science (ICTS) • University of California, Los Angeles — UL1TR001881: UCLA Clinical Translational Science Institute • University of California, San Diego — UL1TR001442: Altman Clinical and Translational Research Institute • University of California, San Francisco — UL1TR001872: UCSF Clinical and Translational Science Institute

## Pending

Arkansas Children’s Hospital — UL1TR003107: UAMS Translational Research Institute • Baylor College of Medicine — None (Voluntary) • Children’s Hospital of Philadelphia — UL1TR001878: Institute for Translational Medicine and Therapeutics • Cincinnati Children’s Hospital Medical Center — UL1TR001425: Center for Clinical and Translational Science and Training • Emory University — UL1TR002378: Georgia Clinical and Translational Science Alliance • HonorHealth — None (Voluntary) • Loyola University Chicago — UL1TR002389: The Institute for Translational Medicine (ITM) • Medical College of Wisconsin — UL1TR001436: Clinical and Translational Science Institute of Southeast Wisconsin • MedStar Health Research Institute — UL1TR001409: The Georgetown-Howard Universities Center for Clinical and Translational Science (GHUCCTS) • MetroHealth — None (Voluntary) • Montana State University — U54GM115371: American Indian/Alaska Native CTR • NYU Langone Medical Center — UL1TR001445: Langone Health’s Clinical and Translational Science Institute • Ochsner Medical Center — U54GM104940: Louisiana Clinical and Translational Science (LA CaTS) Center • Regenstrief Institute — UL1TR002529: Indiana Clinical and Translational Science Institute • Sanford Research — None (Voluntary) • Stanford University — UL1TR003142: Spectrum: The Stanford Center for Clinical and Translational Research and Education • The Rockefeller University — UL1TR001866: Center for Clinical and Translational Science • The Scripps Research Institute — UL1TR002550: Scripps Research Translational Institute • University of Florida — UL1TR001427: UF Clinical and Translational Science Institute • University of New Mexico Health Sciences Center — UL1TR001449: University of New Mexico Clinical and Translational Science Center • University of Texas Health Science Center at San Antonio — UL1TR002645: Institute for Integration of Medicine and Science • Yale New Haven Hospital — UL1TR001863: Yale Center for Clinical Investigation

## Funding

National Institutes of Health grant CD2H NCATS U24 TR002306 (JTR, MH, PNR)

National Institutes of Health grant NHLBI RECOVER Agreement OT2HL161847-01 (JTR, MAH, PNR)

National Institutes of Health grant Office of the Director Monarch Initiative R24 OD011883 (MAH, PNR) National Institutes of Health grant NHGRI Center of Excellence in Genome Sciences RM1 HG010860 (MH, PNR)

Director, Office of Science, Office of Basic Energy Sciences of the U.S. Department of Energy Contract No. DE-AC02-05CH11231 (JTR)

Donald A. Roux Family Fund at the Jackson Laboratory; (PNR)

Marsico Family at the University of Colorado Anschutz (MAH)

## Author contributions

Conceptualization: JTR, PNR

Methodology: JTR, HB, TB, JJL, TC, BL, EC, BC, MG, KW, LC, TF, NA, BA, TMM, GK, JAM, GV, DS, CGC, CM-B, AW, RM, JB,

Investigation: JTR, HB, CA, AES, HD, KK

Funding acquisition: MAH, PNR

Supervision: JTR, MAH, PNR

Writing – original draft: JTR, PNR

Writing – review & editing: JTR, DLA, PRB, JLS, EH

## Data and materials availability

The analyses described in this publication were conducted with data accessed through the NCATS N3C Data Enclave covid.cd2h.org/enclave. Researchers can apply for access to the data as described in https://ncats.nih.gov/n3c/. The code for performing the semantic analysis, clustering, and generalizability assessment is freely available at https://github.com/National-COVID-Cohort-Collaborative/semanticsimilarity and https://github.com/National-COVID-Cohort-Collaborative/kernelkm. Additional code for defining the cohort and transforming raw OMOP data for this analysis is available through the NCATS N3C Data Enclave covid.cd2h.org/enclave with access procedures as described above. The project is available under the Data Use Request RP-5677B5 “Characterization of long-COVID: definition, stratification, and multi-modal analysis”.

