## Supplementary material for "Generalizable Long COVID Subtypes: Findings from the NIH N3C and RECOVER Programs": Online Supplemental Material

**Table S1. Neuropsychiatric** HPO terms that were observed (✓) or not (-) in the EHR data.

| | Term | observed? |
|---|---|---|
| 1 | Aphasia (HP:0002381) | ✓ |
| 2 | Slurred speech (HP:0001350) | ✓ |
| 3 | Agnosia (HP:0010524) | ✓ |
| 4 | Confusion (HP:0001289) | ✓ |
| 5 | Encephalopathy (HP:0001298) | ✓ |
| 6 | Headache (HP:0002315) | ✓ |
| 7 | Migraine (HP:0002076) | ✓ |
| 8 | Anosmia (HP:0000458) | ✓ |
| 9 | Parageusia (HP:0031249) | ✓ |
| 10 | Depression (HP:0000716) | ✓ |
| 11 | Memory impairment (HP:0002354) | ✓ |
| 12 | Attention deficit hyperactivity disorder (HP:0007018) | ✓ |
| 13 | Auditory hallucinations (HP:0008765) | ✓ |
| 14 | Hallucinations (HP:0000738) | ✓ |
| 15 | Visual hallucinations (HP:0002367) | ✓ |
| 16 | Abnormal reflex (HP:0031826) | ✓ |
| 17 | Abnormality of movement (HP:0100022) | ✓ |
| 18 | Ataxia (HP:0001251) | ✓ |
| 19 | Dysarthria (HP:0001260) | ✓ |
| 20 | Dysphagia (HP:0002015) | ✓ |
| 21 | Dystonia (HP:0001332) | ✓ |
| 22 | Hyperesthesia (HP:0100963) | ✓ |
| 23 | Hyperkinetic movements (HP:0002487) | ✓ |
| 24 | Muscle weakness (HP:0001324) | ✓ |
| 25 | Orthostatic hypotension (HP:0001278) | ✓ |
| 26 | Paresthesia (HP:0003401) | ✓ |
| 27 | Polyneuropathy (HP:0001271) | ✓ |
| 28 | Seizure (HP:0001250) | ✓ |
| 29 | Skeletal muscle atrophy (HP:0003202) | ✓ |
| 30 | Somatic sensory dysfunction (HP:0003474) | ✓ |
| 31 | Tremor (HP:0001337) | ✓ |
| 32 | Insomnia (HP:0100785) | ✓ |
| 33 | Restless legs (HP:0012452) | ✓ |
| 34 | Sleep apnea (HP:0010535) | ✓ |
| 35 | Anomic aphasia (HP:0030784) | - |
| 36 | Bilingual aphasia (HP:0033849) | - |
| 37 | Expressive aphasia (HP:0002427) | - |
| 38 | Receptive aphasia (HP:0033848) | - |
| 39 | Bradykinesia (HP:0002067) | - |
| 40 | Bradyphrenia (HP:0031843) | - |
| 41 | Cognitive impairment (HP:0100543) | - |
| 42 | Diminished ability to concentrate (HP:0031987) | - |
| 43 | Tachyphrenia (HP:0033844) | - |
| 44 | Ageusia (HP:0041051) | - |
| 45 | Hypogeusia (HP:0000224) | - |
| 46 | Hyposmia (HP:0004409) | - |

Continued on next page

**Table S1 – continued from previous page**

| | Term | observed? |
|---|---|---|
| 47 | Phantageusia (HP:0033847) | - |
| 48 | Phantosmia (HP:0033693) | - |
| 49 | Aggressive behavior (HP:0000718) | - |
| 50 | Dysphoria (HP:0033838) | - |
| 51 | Emotional lability (HP:0000712) | - |
| 52 | Euphoria (HP:0031844) | - |
| 53 | Mania (HP:0100754) | - |
| 54 | Sense of impending doom (HP:0033845) | - |
| 55 | Suicidal ideation (HP:0031589) | - |
| 56 | Tearfulness (HP:0033705) | - |
| 57 | Anterograde memory impairment (HP:0033689) | - |
| 58 | Long term memory impairment (HP:0033688) | - |
| 59 | Procedural memory loss (HP:0033691) | - |
| 60 | Short term memory impairment (HP:0033687) | - |
| 61 | Anxiety (HP:0000739) | - |
| 62 | Apathy (HP:0000741) | - |
| 63 | Brain fog (HP:0033630) | - |
| 64 | Delusions (HP:0000746) | - |
| 65 | Impaired executive functioning (HP:0033051) | - |
| 66 | Impulsivity (HP:0100710) | - |
| 67 | Irritability (HP:0000737) | - |
| 68 | Panic attack (HP:0025269) | - |
| 69 | Phonophobia (HP:0002183) | - |
| 70 | Polydipsia (HP:0001959) | - |
| 71 | Posttraumatic stress symptom (HP:0033676) | - |
| 72 | Short attention span (HP:0000736) | - |
| 73 | Tactile hallucination (HP:0033694) | - |
| 74 | Abnormal exteroceptive sensation (HP:0033747) | - |
| 75 | Babinski sign (HP:0003487) | - |
| 76 | Dysmetria (HP:0001310) | - |
| 77 | Facial paralysis (HP:0007209) | - |
| 78 | Frontal release signs (HP:0000743) | - |
| 79 | Gait disturbance (HP:0001288) | - |
| 80 | Hand muscle weakness (HP:0030237) | - |
| 81 | Hypoesthesia (HP:0033748) | - |
| 82 | Hypotonia (HP:0001252) | - |
| 83 | Muscle spasm (HP:0003394) | - |
| 84 | Parkinsonism (HP:0001300) | - |
| 85 | Rigidity (HP:0002063) | - |
| 86 | Spasticity (HP:0001257) | - |
| 87 | Unilateral facial palsy (HP:0012799) | - |
| 88 | Maintenance insomnia (HP:0031355) | - |
| 89 | Sleep disturbance (HP:0002360) | - |
| 90 | Sleep onset insomnia (HP:0031354) | - |
| 91 | Terminal insomnia (HP:0031356) | - |

**Table S2. ENT** HPO terms that were observed (✓) or not (-) in the EHR data.

| | Term | observed? |
|---|---|---|
| 92 | Nasal congestion (HP:0001742) | ✓ |
| 93 | Pharyngalgia (HP:0033050) | ✓ |
| 94 | Ear pain (HP:0030766) | ✓ |
| 95 | Hearing impairment (HP:0000365) | ✓ |
| 96 | Hyperacusis (HP:0010780) | ✓ |
| 97 | Tinnitus (HP:0000360) | ✓ |
| 98 | Vertigo (HP:0002321) | ✓ |
| 99 | Dysphonia (HP:0001618) | - |
| 100 | Rhinitis (HP:0012384) | - |
| 101 | Pulsatile tinnitus (HP:0008629) | - |

**Table S3. Skin** HPO terms that were observed (✓) or not (-) in the EHR data.

| | Term | observed? |
|---|---|---|
| 102 | Dermatographic urticaria (HP:0011971) | ✓ |
| 103 | Skin rash (HP:0000988) | ✓ |
| 104 | Alopecia (HP:0001596) | - |
| 105 | Flushing (HP:0031284) | - |
| 106 | Fragile nails (HP:0001808) | - |
| 107 | Hyperhidrosis (HP:0000975) | - |
| 108 | Petechiae (HP:0000967) | - |
| 109 | Pruritus (HP:0000989) | - |
| 110 | Pseudo-chilblain (HP:0033696) | - |
| 111 | Scaling skin (HP:0040189) | - |

**Table S4. Pulmonary** HPO terms that were observed (✓) or not (-) in the EHR data.

| | Term | observed? |
|---|---|---|
| 112 | Atelectasis (HP:0100750) | ✓ |
| 113 | Bronchiectasis (HP:0002110) | ✓ |
| 114 | Pulmonary fibrosis (HP:0002206) | ✓ |
| 115 | Hypoxemia (HP:0012418) | ✓ |
| 116 | Pleuritis (HP:0002102) | ✓ |
| 117 | Pulmonary embolism (HP:0002204) | ✓ |
| 118 | Cough (HP:0012735) | ✓ |
| 119 | Dyspnea (HP:0002094) | ✓ |
| 120 | Hemoptysis (HP:0002105) | ✓ |
| 121 | Sneeze (HP:0025095) | ✓ |
| 122 | Wheezing (HP:0030828) | ✓ |
| 123 | Abnormal pulmonary thoracic imaging finding (HP:0031983) | - |
| 124 | Centrilobular ground-glass opacification on pulmonary HRCT (HP:0025180) | - |
| 125 | Crazy-paving pattern (HP:0033659) | - |
| 126 | Interlobular septal thickening (HP:0030879) | - |
| 127 | Parenchymal consolidation (HP:0032177) | - |
| 128 | Pleural thickening (HP:0031944) | - |
| 129 | Pulmonary bulla (HP:0032446) | - |
| 130 | Pulmonary interstitial thickening (HP:0033711) | - |
| 131 | Reticular pattern on pulmonary HRCT (HP:0025390) | - |
| 132 | Solid pulmonary nodule (HP:0033609) | - |
| 133 | Subsolid pulmonary nodule (HP:0033610) | - |
| 134 | Subpleural curvilinear line (HP:0033702) | - |
| 135 | Airway obstruction (HP:0006536) | - |
| 136 | Decreased DLCO (HP:0045051) | - |
| 137 | Decreased maximal oxygen uptake (HP:0033760) | - |
| 138 | Decreased RV/TLC ratio (HP:0033773) | - |
| 139 | Ground-glass opacification (HP:0025179) | - |
| 140 | Oxygen desaturation on exertion (HP:0030874) | - |
| 141 | Reduced FEV1/FVC ratio (HP:0030877) | - |
| 142 | Reduced forced expiratory volume in one second (HP:0032342) | - |
| 143 | Reduced forced vital capacity (HP:0032341) | - |
| 144 | Reduced functional residual capacity (HP:0033750) | - |
| 145 | Reduced residual volume (HP:0033753) | - |
| 146 | Reduced total lung capacity (HP:0033169) | - |
| 147 | Restrictive ventilatory defect (HP:0002091) | - |
| 148 | Exertional dyspnea (HP:0002875) | - |
| 149 | Increased sputum production (HP:0033709) | - |
| 150 | Nonproductive cough (HP:0031246) | - |
| 151 | Pleuritic chest pain (HP:0033771) | - |
| 152 | Productive cough (HP:0031245) | - |
| 153 | Rest dyspnea (HP:0033710) | - |
| 154 | Rhinorrhea (HP:0031417) | - |
| 155 | Rhonchi (HP:0030831) | - |
| 156 | Tachypnea (HP:0002789) | - |

**Table S5. Endocrine** HPO terms that were observed (✓) or not (-) in the EHR data.

| | Term | observed? |
|---|---|---|
| 157 | Diabetes mellitus (HP:0000819) | ✓ |
| 158 | Edema (HP:0000969) | ✓ |
| 159 | Fever (HP:0001945) | ✓ |
| 160 | Hypothermia (HP:0002045) | ✓ |
| 161 | Irregular menstruation (HP:0000858) | ✓ |
| 162 | Menorrhagia (HP:0000132) | ✓ |
| 163 | Renal insufficiency (HP:0000083) | ✓ |
| 164 | Urinary incontinence (HP:0000020) | ✓ |
| 165 | Decreased glomerular filtration rate (HP:0012213) | - |
| 166 | Female sexual dysfunction (HP:0030014) | - |
| 167 | Heat intolerance (HP:0002046) | - |
| 168 | Low-grade fever (HP:0011134) | - |
| 169 | Male sexual dysfunction (HP:0040307) | - |
| 170 | Pancreatitis (HP:0001733) | - |
| 171 | Postmenopausal bleeding (HP:0033840) | - |
| 172 | Recurrent fever (HP:0001954) | - |
| 173 | Temperature instability (HP:0005968) | - |
| 174 | Testicular pain (HP:0033839) | - |

**Table S6. Immunology** HPO terms that were observed (✓) or not (-) in the EHR data.

| | Term | observed? |
|---|---|---|
| 175 | Anaphylactic shock (HP:0100845) | ✓ |
| 176 | Lymphadenopathy (HP:0002716) | ✓ |
| 177 | Lymphopenia (HP:0001888) | ✓ |
| 178 | Antinuclear antibody positivity (HP:0003493) | - |
| 179 | Anti-thyroid peroxidase antibody positivity (HP:0025379) | - |
| 180 | Anti-thyroglobulin antibody positivity (HP:0032069) | - |

**Table S7. General** HPO terms that were observed (✓) or not (-) in the EHR data.

| | Term | observed? |
|---|---|---|
| 181 | Arthritis (HP:0001369) | ✓ |
| 182 | Asthenia (HP:0025406) | ✓ |
| 183 | Difficulty walking (HP:0002355) | ✓ |
| 184 | Fatigue (HP:0012378) | ✓ |
| 185 | Shivering (HP:0025144) | ✓ |
| 186 | Xerostomia (HP:0000217) | ✓ |
| 187 | Arthralgia (HP:0002829) | ✓ |
| 188 | Chest pain (HP:0100749) | ✓ |
| 189 | Limb pain (HP:0009763) | ✓ |
| 190 | Myalgia (HP:0003326) | ✓ |
| 191 | Pain (HP:0012531) | ✓ |
| 192 | Chest tightness (HP:0031352) | - |
| 193 | Chills (HP:0025143) | - |
| 194 | Coldness (HP:0033850) | - |
| 195 | Diminished health-related quality of life (HP:0033665) | - |
| 196 | Diminished mental health (HP:0033667) | - |
| 197 | Diminished physical functioning (HP:0033666) | - |
| 198 | Exercise intolerance (HP:0003546) | - |
| 199 | Frailty (HP:0033675) | - |
| 200 | Impaired ability to bathe oneself (HP:0031059) | - |
| 201 | Impaired ability to dress oneself (HP:0031060) | - |
| 202 | Impairment of activities of daily living (HP:0031058) | - |
| 203 | Malaise (HP:0033834) | - |
| 204 | Night sweats (HP:0030166) | - |
| 205 | Occupational disability (HP:0033695) | - |
| 206 | Postexertional malaise (HP:0030973) | - |
| 207 | Stiff neck (HP:0025258) | - |
| 208 | Weight loss (HP:0001824) | - |
| 209 | Body ache (HP:0033047) | - |
| 210 | Bone pain (HP:0002653) | - |
| 211 | Intrascapular pain (HP:0033746) | - |
| 212 | Neuralgia (HP:0033345) | - |

**Table S8. Cardiovascular** HPO terms that were observed (✓) or not (-) in the EHR data.

| | Term | observed? |
|---|---|---|
| 213 | Bradycardia (HP:0001662) | ✓ |
| 214 | Hypotension (HP:0002615) | ✓ |
| 215 | Myocarditis (HP:0012819) | ✓ |
| 216 | Tachycardia (HP:0001649) | ✓ |
| 217 | Angina pectoris (HP:0001681) | ✓ |
| 218 | Palpitations (HP:0001962) | ✓ |
| 219 | Elevated myocardial native T1 (HP:4000006) | - |
| 220 | Elevated myocardial native T2 (HP:4000003) | - |
| 221 | Hypertension (HP:0000822) | - |
| 222 | Increased circulating troponin T concentration (HP:0410174) | - |
| 223 | Increased circulating troponin I concentration (HP:0410173) | - |
| 224 | Increased heart rate variability (HP:0031862) | - |
| 225 | Increased left ventricular end-diastolic volume (HP:0033755) | - |
| 226 | Myocardial late gadolinium enhancement (HP:4000004) | - |
| 227 | Pericardial effusion (HP:0001698) | - |
| 228 | Pericardial late gadolinium enhancement (HP:4000005) | - |
| 229 | Reduced ejection fraction (HP:0012664) | - |
| 230 | Venous thrombosis (HP:0004936) | - |
| 231 | Stroke (HP:0001297) | - |
| 232 | Syncope (HP:0001279) | - |

**Table S9. Gastrointestinal** HPO terms that were observed (✓) or not (-) in the EHR data.

| | Term | observed? |
|---|---|---|
| 233 | Gastroesophageal reflux (HP:0002020) | ✓ |
| 234 | Gastroparesis (HP:0002578) | ✓ |
| 235 | Hepatic steatosis (HP:0001397) | ✓ |
| 236 | Hepatomegaly (HP:0002240) | ✓ |
| 237 | Hepatitis (HP:0012115) | ✓ |
| 238 | Malnutrition (HP:0004395) | ✓ |
| 239 | Splenomegaly (HP:0001744) | ✓ |
| 240 | Abdominal pain (HP:0002027) | ✓ |
| 241 | Bowel incontinence (HP:0002607) | ✓ |
| 242 | Constipation (HP:0002019) | ✓ |
| 243 | Diarrhea (HP:0002014) | ✓ |
| 244 | Nausea (HP:0002018) | ✓ |
| 245 | Vomiting (HP:0002013) | ✓ |
| 246 | Gastric ulcer (HP:0002592) | - |
| 247 | Pancreatic steatosis (HP:0033757) | - |
| 248 | Abdominal symptom (HP:0011458) | - |
| 249 | Anorexia (HP:0002039) | - |
| 250 | Early satiety (HP:0033842) | - |
| 251 | Poor appetite (HP:0004396) | - |

**Table S10.** **Laboratory** HPO terms that were observed (✓) or not (-) in the EHR data.

| | **Term** | **observed?** |
|---|---|---|
| 252 | Decreased circulating calcifediol concentration (HP:0012053) | ✓ |
| 253 | Elevated circulating alkaline phosphatase concentration (HP:0003155) | ✓ |
| 254 | Elevated circulating alanine aminotransferase concentration (HP:0031964) | ✓ |
| 255 | Elevated circulating aspartate aminotransferase concentration (HP:0031956) | ✓ |
| 256 | Elevated circulating creatinine concentration (HP:0003259) | ✓ |
| 257 | Elevated circulating creatine kinase concentration (HP:0003236) | ✓ |
| 258 | Elevated circulating thyroid-stimulating hormone concentration (HP:0002925) | ✓ |
| 259 | Elevated erythrocyte sedimentation rate (HP:0003565) | ✓ |
| 260 | Elevated gamma-glutamyltransferase level (HP:0030948) | ✓ |
| 261 | Hypocalcemia (HP:0002901) | ✓ |
| 262 | Hypofibrinogenemia (HP:0011900) | ✓ |
| 263 | Hyperglycemia (HP:0003074) | ✓ |
| 264 | Hypoglycemia (HP:0001943) | ✓ |
| 265 | Hypophosphatemia (HP:0002148) | ✓ |
| 266 | Increased circulating ferritin concentration (HP:0003281) | ✓ |
| 267 | Thrombocytopenia (HP:0001873) | ✓ |
| 268 | Elevated circulating C-reactive protein concentration (HP:0011227) | - |
| 269 | Elevated circulating D-dimer concentration (HP:0033106) | - |
| 270 | Elevated circulating soluble CD25 concentration (HP:0033833) | - |
| 271 | Increased circulating interleukin 6 (HP:0030783) | - |
| 272 | Increased circulating lactate dehydrogenase concentration (HP:0025435) | - |
| 273 | Increased circulating NT-proBNP concentration (HP:0031185) | - |
| 274 | Increased circulating procalcitonin concentration (HP:0032308) | - |

**Table S11. Eye** HPO terms that were observed (✓) or not (-) in the EHR data.

| | Term | observed? |
|---|---|---|
| 275 | Conjunctivitis (HP:0000509) | ✓ |
| 276 | Diplopia (HP:0000651) | ✓ |
| 277 | Keratoconjunctivitis sicca (HP:0001097) | ✓ |
| 278 | Ocular pain (HP:0200026) | ✓ |
| 279 | Vitreous floaters (HP:0100832) | ✓ |
| 280 | Blindness (HP:0000618) | - |
| 281 | Blurred vision (HP:0000622) | - |
| 282 | Gaze-evoked nystagmus (HP:0000640) | - |
| 283 | Ocular pruritus (HP:0033841) | - |
| 284 | Peripheral visual field loss (HP:0007994) | - |
| 285 | Photophobia (HP:0000613) | - |
| 286 | Red eye (HP:0025337) | - |
| 287 | Visual loss (HP:0000572) | - |

| Covariate | Sig.? |
|---|---|
| AKI | ✓ |
| Chronic lung disease | ✓ |
| Congestive heart failure | ✓ |
| Coronary artery disease | ✓ |
| Depression | ✓ |
| Diabetes (complicated) | ✓ |
| Diabetes (uncomplicated) | ✓ |
| Heart failure | ✓ |
| Hypertension | ✓ |
| Kidney disease | ✓ |
| Obsesity | ✓ |
| Peripheral vascular disease | ✓ |
| Systemic corticosteroids | ✓ |
| At least one vaccine recorded | - |
| Cardiomyopathies | - |
| Cerebrovascular disease | - |
| Dementia | - |
| Down syndrome | - |
| Hemiplegia or paraplegia | - |
| HIV infection | - |
| Malignant cancer | - |
| Metastatic solid tumor cancer | - |
| Mild liver disease | - |
| Moderate liver disease | - |
| Multiple sclerosis | - |
| Myocardial infarction | - |
| Obstructive sleep apnea | - |
| Peptic ulcer | - |
| Psychosis | - |
| Sickle cell disease | - |
| Substance abuse | - |
| Thalassemia | - |
| Tobacco smoker | - |
| Tuberculosis | - |
| Rheumatoligical disease | - |

**Table S12. Covariates assessed prior to acute COVID-19.** The statistically significant covariates are shown in the main manuscript. n.s.: non-significant. ✓: statistically sigificant with Bonferroni-corrected $p < 0.05$. Abbreviations: AKI: acute kidney injury. IMV: Invasive mechanical ventilation.

| Covariate | Sig.? |
|---|---|
| AKI | ✓ |
| IMV | ✓ |
| COVID regimen corticosteroids | ✓ |
| Sepsis | ✓ |
| Vasopressors | ✓ |
| Remdesivir | ✓ |
| Death during COVID hospitalization | - |
| ECMO | - |
| COVID diagnosis during hospitalization | - |

**Table S13. Covariates assessed during acute COVID-19.** The statistically significant covariates are shown in the main manuscript. n.s.: non-significant. ✓: statistically sigificant with Bonferroni-corrected $p < 0.05$. Abbreviations: AKI: acute kidney injury. IMV: Invasive mechanical ventilation.

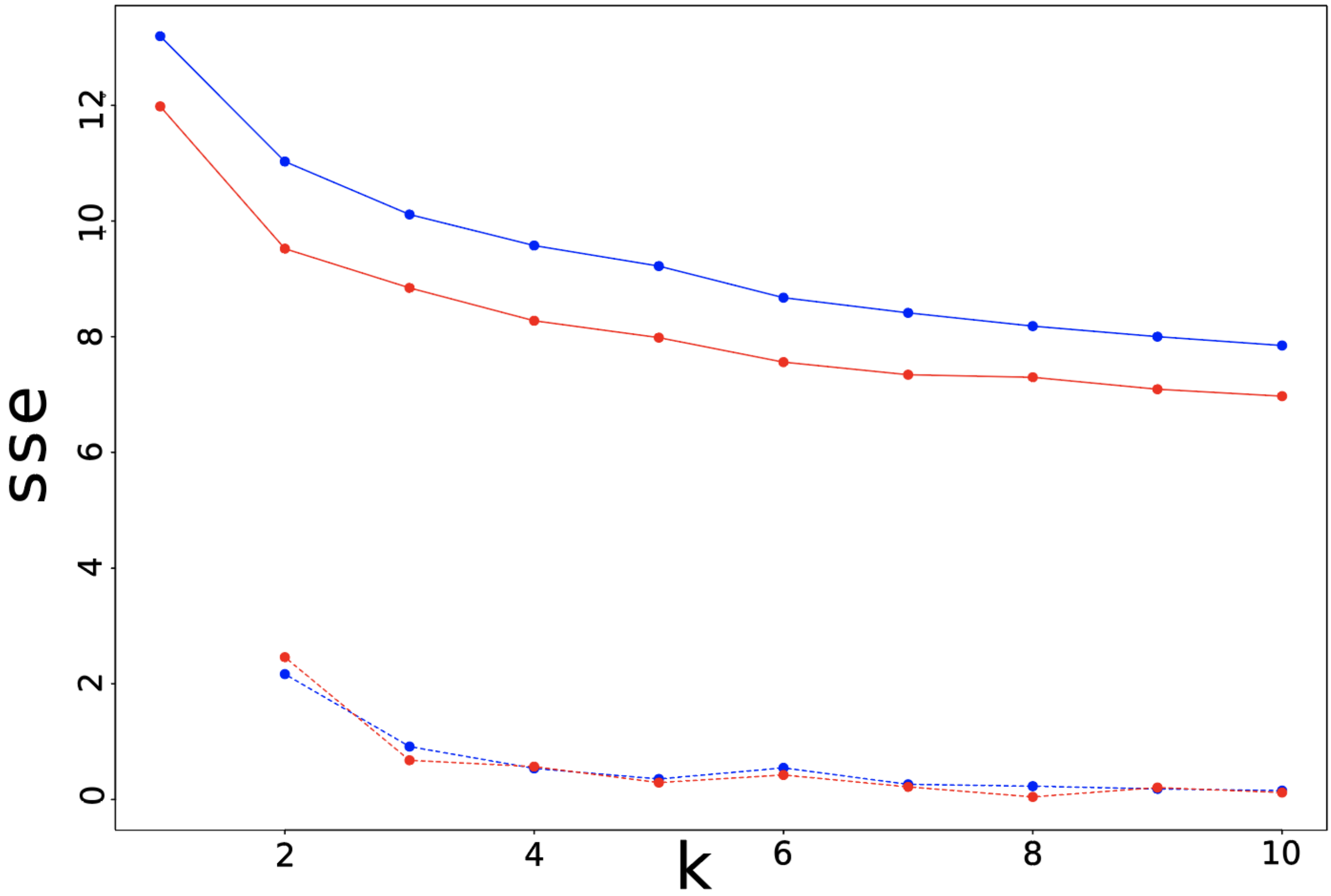


**Figure S1. Determination of optimal k using the elbow method.** For patients from data partner 1 (blue) or data partners 2 - 5 (red), patients were clustered with the k-means algorithm using values of k shown on the x-axis. For each value of k, the sum of squared error (SSE) (x $10^4$) of the distance from each patient to the centroid of the patient's cluster (solid lines) and the decrease in SSE (x $10^4$) from the previous k (dotted lines) is shown on the y-axis.

| | HPO | Cluster 1 | Cluster 2 | Cluster 3 | Cluster 4 |
|---|---|---|---|---|---|
| | Vertigo | 28.4% | 8.2% | 7.7% | 18.1% |
| | Hypotension | 21.8% | 16.0% | 1.3% | 2.4% |
| | Palpitations | 22.6% | 2.4% | 3.0% | 15.5% |
| | Tachycardia | 49.4% | 17.4% | 3.0% | 16.0% |
| | Fever | 40.6% | 19.5% | 5.4% | 10.8% |
| | Diarrhea | 38.3% | 15.4% | 4.7% | 5.2% |
| | Nausea | 24.5% | 7.2% | 3.4% | 6.8% |
| | Asthenia | 39.5% | 18.4% | 2.0% | 8.4% |
| | Chest pain | 49.8% | 8.9% | 0.0% | 49.3% |
| | Fatigue | 64.8% | 23.5% | 4.0% | 56.4% |
| | Myalgia | 29.1% | 3.8% | 0.0% | 20.2% |
| | Pain | 77.8% | 19.8% | 2.3% | 72.7% |
| | Lymphopenia | 46.7% | 61.8% | 2.7% | 4.2% |
| | Elevated ALP | 25.7% | 21.8% | 1.3% | 1.0% |
| | Elevated ASAT | 46.7% | 53.6% | 0.7% | 1.3% |
| | Elevated creatinine | 29.1% | 38.9% | 2.3% | 1.8% |
| | Hyperglycemia | 77.4% | 78.8% | 20.5% | 26.8% |
| | Hypocalcemia | 62.5% | 72.4% | 1.0% | 3.4% |
| | Increased ferritin | 18.4% | 20.1% | 0.0% | 0.3% |
| | Thrombocytopenia | 30.7% | 35.8% | 1.7% | 1.6% |
| | Abnormality of movement | 20.7% | 3.8% | 7.7% | 7.6% |
| | Depression | 21.8% | 8.9% | 8.7% | 7.3% |
| | Headache | 36.8% | 6.5% | 14.1% | 23.4% |
| | Insomnia | 23.0% | 8.5% | 8.1% | 8.7% |
| | Cough | 59.8% | 25.3% | 44.0% | 36.7% |
| | Hypoxemia | 69.7% | 64.5% | 17.8% | 17.3% |

**Figure S2. Summary of phenotypic feature distribution in the four clusters.** HPO terms are shown if Pearson's chi-squared test on the numbers of patients in each category with the feature was significant with $p < 0.00001$ and if at least 20% of patients in at least one cluster had the feature. Terms are grouped in categories shown color coded on the left.

| | HPO | Cluster 1 | Cluster 2 | Cluster 3 | Cluster 4 | Cluster 5 |
|---|---|---|---|---|---|---|
| | Vertigo | 27.6% | 8.2% | 19.1% | 19.7% | 2.2% |
| | Hypotension | 22.4% | 16.1% | 1.9% | 2.6% | 1.7% |
| | Palpitations | 21.7% | 2.5% | 3.7% | 19.7% | 2.2% |
| | Tachycardia | 47.6% | 17.9% | 4.3% | 20.7% | 3.5% |
| | Fever | 39.8% | 20.4% | 2.5% | 12.9% | 7.9% |
| | Diarrhea | 38.6% | 15.8% | 9.3% | 6.1% | 1.3% |
| | Nausea | 24.4% | 7.9% | 7.4% | 7.1% | 1.3% |
| | Asthenia | 40.9% | 19.4% | 6.2% | 6.1% | 3.5% |
| | Chest pain | 48.4% | 9.3% | 4.3% | 60.5% | 0.4% |
| | Fatigue | 64.6% | 24.4% | 25.3% | 50.5% | 15.7% |
| | Myalgia | 29.1% | 3.9% | 3.1% | 23.9% | 0.0% |
| | Pain | 77.2% | 20.1% | 14.8% | 85.1% | 2.6% |
| | Lymphopenia | 48.0% | 62.7% | 5.6% | 4.2% | 3.5% |
| | Elevated ALP | 25.6% | 22.2% | 1.2% | 1.9% | 1.7% |
| | Elevated ASAT | 48.0% | 55.6% | 0.6% | 1.6% | 1.3% |
| | Elevated creatinine | 30.3% | 40.5% | 4.3% | 1.3% | 1.3% |
| | Hyperglycemia | 79.1% | 77.8% | 16.0% | 27.8% | 28.8% |
| | Hypocalcemia | 63.4% | 74.9% | 1.2% | 4.9% | 1.7% |
| | Increased ferritin | 18.9% | 21.1% | 0.0% | 0.3% | 0.0% |
| | Thrombocytopenia | 31.1% | 37.3% | 4.9% | 1.3% | 0.4% |
| | Abnormality of movement | 20.9% | 3.2% | 19.1% | 6.5% | 1.7% |
| | Depression | 23.2% | 8.2% | 26.5% | 3.9% | 0.0% |
| | Headache | 37.4% | 6.1% | 32.7% | 22.0% | 5.7% |
| | Insomnia | 24.0% | 8.6% | 7.4% | 8.7% | 7.9% |
| | Cough | 59.1% | 24.7% | 15.4% | 38.8% | 59.8% |
| | Hypoxemia | 70.5% | 64.5% | 9.3% | 18.8% | 25.3% |

**Figure S3. Summary of phenotypic feature distribution in the five clusters.** HPO terms are shown if Pearson's chi-squared test on the numbers of patients in each category with the feature was significant with $p < 0.00001$ and if at least 20% of patients in at least one cluster had the feature. Terms are grouped in categories shown color coded on the left.
